# Short-term and Long-Term Healthcare Costs Attributable to diagnosed COVID-19 in Ontario; Canada: A Population-Based Matched Cohort Study

**DOI:** 10.1101/2024.09.04.24313064

**Authors:** Beate Sander, Sharmistha Mishra, Sarah Swayze, Yeva Sahakyan, Raquel Duchen, Kieran Quinn, Naveed Janjua, Hind Sbihi, Jeffrey Kwong

## Abstract

**Objectives:** Estimates of health system costs due to COVID-19, especially for long-term disability (post COVID-19 condition [PCC]) are key to health system planning, but attributable cost data remain scarce. We characterized COVID-19-attributable costs from the health system perspective.

**Methods:** Population-based matched cohort study in Ontario, Canada, using health administrative data. To assign attribution to COVID-19, individuals, defined as exposed (positive SARS-CoV-2 PCR test, 01/2020-12/2020) were matched 1:1 to an unexposed individuals (01/2016-12/2018). Historical matching was used to reduce biases due to overall reductions in healthcare during the pandemic and contamination bias. The index date was defined as the first occurrence of positive SARS-CoV-2 PCR test. We used phase-of-care costing to calculate mean attributable per-person costs (2023 CAD), standardized to 10 days, during four phases of illness: pre-index date, acute care, post-acute care (suggestive of PCC), and terminal phase (stratified by early and late deaths). Finally, we estimated total costs at 360 days by combining costs with survival estimates.

**Results:** Of 165,838 exposed individuals, 159,817 were matched (mean age 40±20 years, 51% female). Mean (95%CI) attributable 10-day costs per person were $1 ($-4, $6) pre-index, $240 ($231, $249) during acute care, and $18 ($14, $21) during post-acute phases. During the terminal phase, mean attributable costs were $3,928 ($3,471, $4,384) for early deaths and $1,781 ($1,182, $2,380) for late deaths. Hospitalizations accounted for 42% to 100% of total costs. Compared to males, costs among females were lower during the acute care phase, but higher during the post-acute care phase. Mean cumulative per-person cost at 360 days was $2,553 ($2,348, $2,756); females had lower costs ($2,194 [$1,945, $2,446]) than males ($2,921 [$2,602, $3,241]).

**Conclusions:** SARS-CoV-2 infection is associated with substantial long-term healthcare costs, consistent with our understanding of the PCC. Understanding phase-specific costs can inform health sector budget planning, future economic evaluations, and pandemic planning.

## Background

Although only recently emerged, SARS-CoV-2 has quickly become a leading burden to the Canadian healthcare system [1]. The health impact of SARS-CoV-2 ranges from asymptomatic to long-term disability due to the post COVID-19 condition (PCC), with the potential to cause substantial long-term costs to health systems [2, 3]. PCC is characterized by the continuation or occurrence of new symptoms three months following the initial SARS-CoV-2 infection, with these symptoms persisting for at least two months without any other explanation [4]. The US Household Pulse Survey reported that 31% of US adults with a history of SARS-COV-2 infection experienced PCC, and the Canadian COVID-19 Antibody and Health Survey reported a 15% prevalence of PCC among Canadian adults with a history of SARS-CoV-2 infection [5, 6]. Emerging data suggest an association between PCC and increased health service utilization for individuals requiring both outpatient and inpatient care [3, 7–9]. A US study found that over a period of seven months, overall healthcare costs nearly doubled for patients with PCC compared to those without it, with costs anticipated to rise further with extended follow-up [10]. Given the high prevalence of people who have been infected with SARS-CoV-2 (83% of Ontario adults by January 2024 [11]), evidence of long-term effects of COVID-19 and its impact on health services utilization is vital for health systems to understand the costs of caring for individuals with PCC across different sectors of the health system.

Previous studies that sought to characterize costs associated with COVID-19 were either focused on inpatient costs [12], the acute period [13], or the post-acute period only [14, 15]. These studies potentially underestimate the overall financial impact of COVID-19 as they did not comprehensively assess the healthcare costs beyond these periods. The importance of considering a longer time horizon is evident in recent studies documenting the long-term health [2, 3] and resource use [7, 8] impact of the PCC. Understanding the economic burden of COVID-19 at different stages of a disease allows for effective resource allocation and planning tailored to each phase.

The purpose of our study was to characterize acute and long-term COVID-19-attributable healthcare costs from the Ontario health system perspective among individuals who tested positive for SARS-CoV-2 compared to matched unexposed individuals, using a phase-of-care approach.

## Methods

We report our study following the RECORD statement for observational studies [16].

### Study design, setting, and population

We conducted an incidence-based matched cohort study to measure the attributable healthcare costs associated with COVID-19 among people registered in the publicly funded Ontario Health Insurance Plan (OHIP). Ontario is Canada’s most populous province, with 40% of the country’s population, and OHIP covers most residents. We used population-based, health administrative datasets that contain information about all publicly funded healthcare encounters covered by OHIP. COVID-19-related information included data on all COVID-19 clinical encounters, publicly funded polymerase chain reaction (PCR) tests, vaccinations provided, and public health case and contact management records. Area-based demographic information such as neighbourhood income quintile was obtained from Census data. All these datasets were linked using unique encoded identifiers and analyzed at ICES, an independent, non-profit research institute. **Table S1**, contains a description of all datasets used in this study.

#### Exposed individuals

We identified individuals as exposed if they tested positive for SARS-CoV-2 by polymerase chain reaction (PCR) between January 1, 2020 and December 31, 2020. This time period allowed for one year of follow-up of individuals classified as exposed, while limiting potential biases due to the emergence of variants of concern in December 2020 and vaccination starting in December 2020 [17]. Index date was defined as the first occurrence of a positive SARS-CoV-2 PCR test for an individual, based on the earliest of symptom onset date recorded in Ontario’s Public Health Case and Contact Management Solution (CCM), specimen collection date, observation date or reporting date recorded in the Ontario Laboratories Information System (OLIS), or case report date in CCM. We excluded individuals with hospital-acquired SARS-CoV-2 infection, defined as follows: those who tested positive on day eight or later after admission, or up to two days after discharge, given a length of stay (LOS) of at least 8 days, or those who were readmitted with a positive test within eight days after discharge from the hospital [18–20].

#### Unexposed individuals

We identified unexposed individuals by taking a 50% random sample of the OHIP-registered population between January 1, 2016, and December 31, 2018. We chose a historical unexposed cohort because: (1) health service use declined by 27%-43% during the pandemic compared to pre-pandemic years, due to system, structural, and behavioural factors [21–23]; and (2) to avoid potential contamination bias because not all individuals with SARS-CoV-2 infection would have had a PCR test for a variety of reasons, such as being ineligible for PCR testing, lacking access to or desire for testing, or being asymptomatic, which ranged from 20-62% [22, 24, 25]. Unexposed individuals were assigned a pseudo-index date at random. Individuals with death dates prior to their pseudo-index dates were excluded from the study. Individuals assigned to the historical unexposed cohort were excluded from the exposed cohort.

Exclusion criteria applying to both groups included: not residing in Ontario, having invalid or missing sex, birthdate, or income quintile information, being aged ≥65 years and not having any healthcare system interactions for the three years prior to the study start date (January 1, 2020, for the exposed group; January 1, 2016, for the unexposed group), being younger than 65 years and not having any healthcare system interactions for the ten years prior to the study start date, being older than 110 years, and residing in long-term care. All individuals were followed for one year from their index/pseudo-index date.

#### Matching

We used a combination of hard and propensity score matching on selected baseline covariates. Each exposed individual was matched to one unexposed individual using nearest-neighbour matching without replacement on index date (day and month) +/-60 days, sex, age category (10-year range for adults, 2–7-year ranges for children and youth), resource utilization band, with a 2-year look-back window, and the logit of the propensity score using a caliper distance of 0.2 standard deviations [26, 27]. Resource utilization band were derived from the Johns Hopkins Adjusted Clinical Groups® (ACGs) System Version 11, and categorizes comorbidity into six groupings from non-utilizer to high complexity of illness [28, 29].

The propensity score included the following baseline measures: public health unit (PHU) of residence, rural/urban residence, census tract of residence (which are not contiguous with the boundaries of local public health units), frailty, high-risk occupation neighbourhood concentration quintile, immigrant status (yes/no), and Ontario Marginalization Index [30] quintiles for age and labour force, material resources, racialized and newcomer populations, and households and dwellings. Frailty was defined using Johns Hopkins ACGs [31]. High-risk occupation neighbourhood concentration quintile was based on the proportion of residents in a census dissemination area (DA) employed as essential workers in the following industries: Sales and Service, Trades, Transport, Equipment Operation, Manufacturing and Utilities, Resources, Agriculture, and Production.

Since healthcare utilization and associated costs tend to increase before death [32], we examined attributable cost prior to death by re-matching exposed individuals who died to unexposed individuals who also died during the observation period on their death dates (day and month) +/-30 days, sex, income quintile, recent immigrant and the logit of the propensity score using a caliper distance of 0.2 standard deviations [26, 27]. The propensity score included the following baseline measures: PHU of residence, rural/urban residence, census tract of residence (not contiguous with the boundaries of the PHUs), high-risk occupation neighbourhood concentration quintile, and Ontario Marginalization Index quintiles [30] for age and labor force, material resources, racialized and newcomer populations, and households and dwellings. Balance for all matches was assessed using standardized differences with a threshold of 0.10 [26, 27].

### Outcomes

The outcome was healthcare costs adjusted for survival over a one-year period following the index date. Costs in 2023 Canadian Dollars were calculated from the Ontario health system perspective (i.e., OHIP), and included all publicly funded health services: inpatient hospitalizations, outpatient hospital visits (same-day surgeries, outpatient cancer therapies and dialyses), emergency department visits, publicly funded drugs (for everyone aged ≥65 years and select younger individuals based on means) physician services, rehabilitation services, complex care, homecare, long-term care, and other (e.g., laboratory tests/services, OHIP-covered non-physician services, assistive devices).

### Analyses

To calculate survival-adjusted costs, we first estimated healthcare costs, standardized to 10 days, by dividing each patient’s total cost by the number of follow-up days, from the index date to the end of follow-up or death and multiplying by 10, using ICES person-level costing methods [33]. Next, we used the phase-of-care costing approach to assign 10-day costs to phases of care along the natural history of disease trajectory [34]. Phase-of-care lengths were determined based on clinical expertise (JK, SM), WHO’s definition of the PCC [4] and joinpoint analysis, identifying the best-fitting points where statistically significant changes occurred in the trend of mean 10-day costs [35]. We defined four phases of care: pre-diagnosis (30 days prior to index date), acute care (80 days post index date), post-acute care (time between acute care phase and terminal phase follow-up), and terminal (60 days prior to death date) phase. We further stratified deaths as “early”, if death occurred within 60 days of index date, and “late” if death occurred more than 60 days following index date. We calculated mean attributable phase-of-care-specific costs (standardized to 10 days) and 95% confidence intervals (CIs) using a generalized estimating equation (GEE) model. Lastly, we combined attributable phase-of-care-specific costs with crude survival data to determine 1-year costs adjusted for survival as described by Yabroff et al [34].

We stratified analyses by age (<2 years, 2-4 years, 5-11 years, 12-17 years, 18-29 years, 30-49 years, 50-69 years, and ≥70 years), sex, income quintile, and resource utilization band quintile.

To understand the impact of phase length on COVID-19-attributable costs, we conducted sensitivity analyses where we varied the lengths of the pre-diagnosis (120 days prior to index date), acute care (30 days post index date), post-acute care (time between acute care phase and terminal phase follow-up) phases.

We used SAS Enterprise Guide 7.15 (SAS Institute, Inc.) for all statistical analyses.

## Results

### Study cohort

Between January 1, 2020, and December 31, 2020, we identified 181,979 Ontario residents who tested positive for SARS-CoV-2 (**Figure 1**). Among them, 165,838 met eligibility criteria for the exposed cohort with a mean age of 40.4 ± 19.7 years and 50.7% were female. There were 6,641,074 unexposed individuals residing in Ontario between January 1, 2016, and December 31, 2018 who were eligible for matching, with a mean age of 40.5 ± 22.8 years and 50.6% were female (**Table 1**). Prior to matching, compared to unexposed individuals, exposed individuals were more likely to be immigrants, and live in urban areas and neighbourhoods with a higher proportion of the population having lower material resources, a higher proportion of racialized and newcomer populations, a higher proportion of essential workers, and lower household incomes.

**Figure 1.**
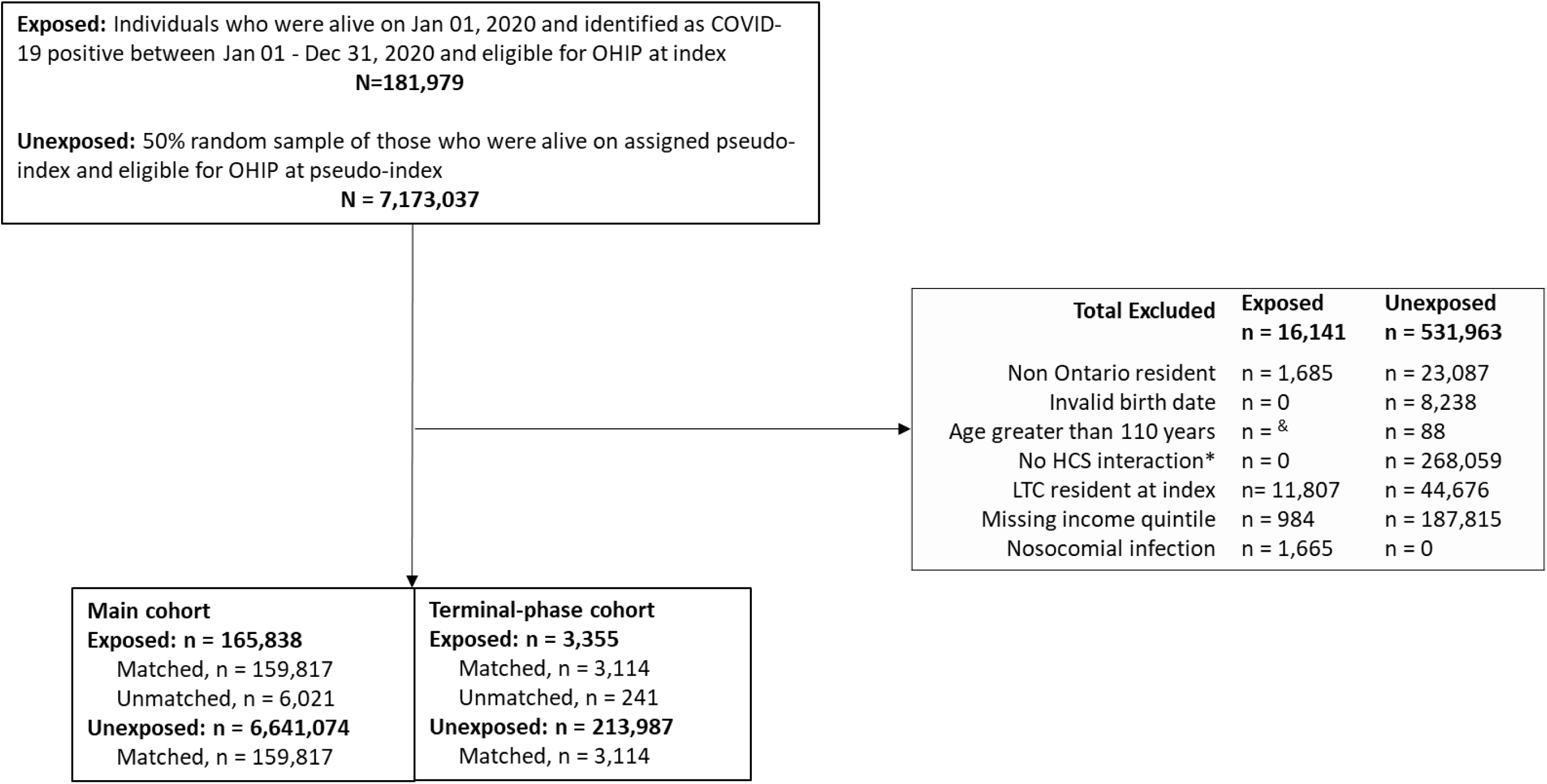
Cohort creation flowchart. HCS: healthcare system interaction; LTC: long-term care ^&^ Due to small cell number this category has been combined with the “Non Ontario resident” category *No interaction with healthcare system was defined as being aged ≥65 years without healthcare system interaction for the three years prior to the study start date or being <65 years without healthcare system interaction for the 10 years prior to the study start date

**Table 1.**
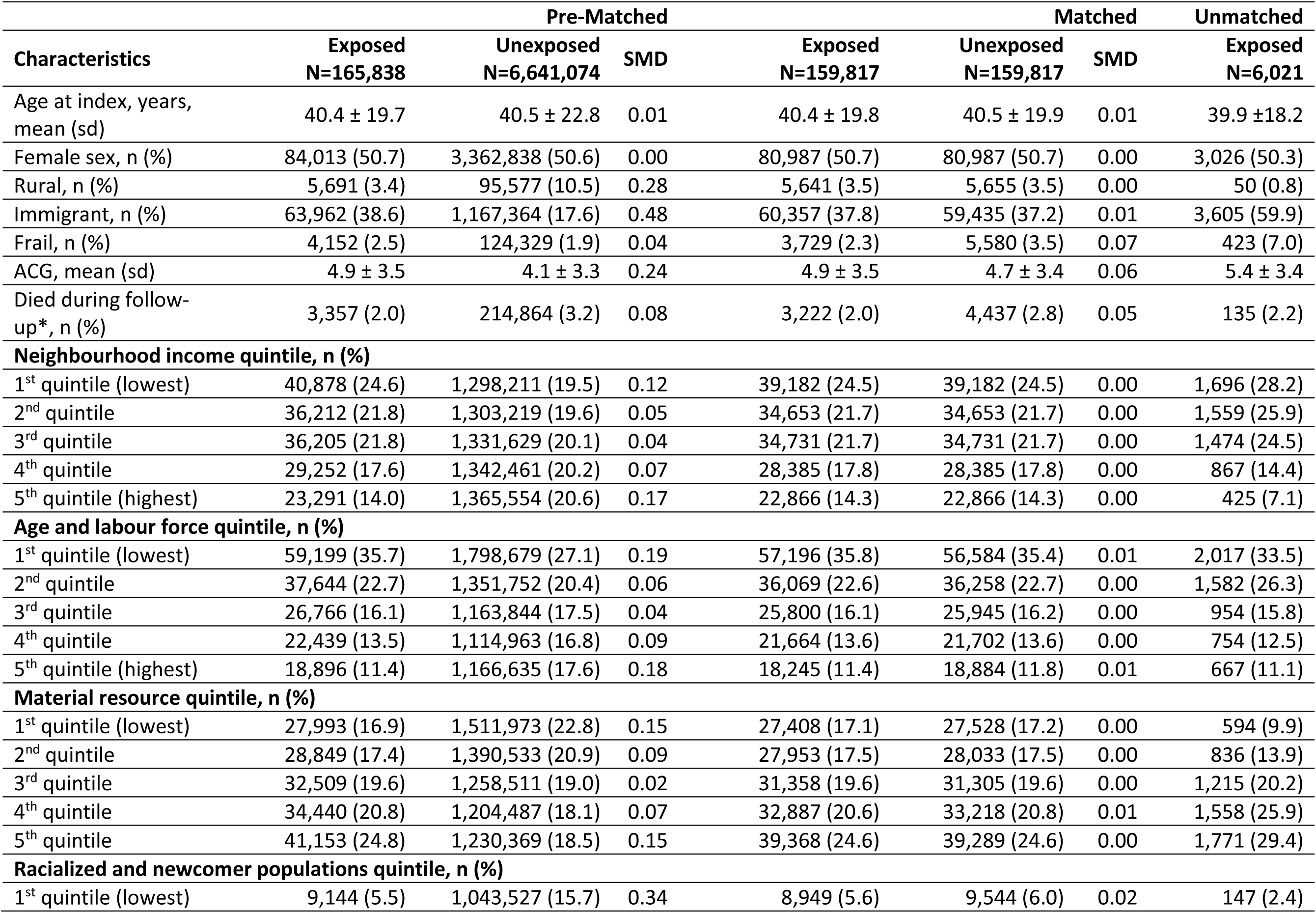
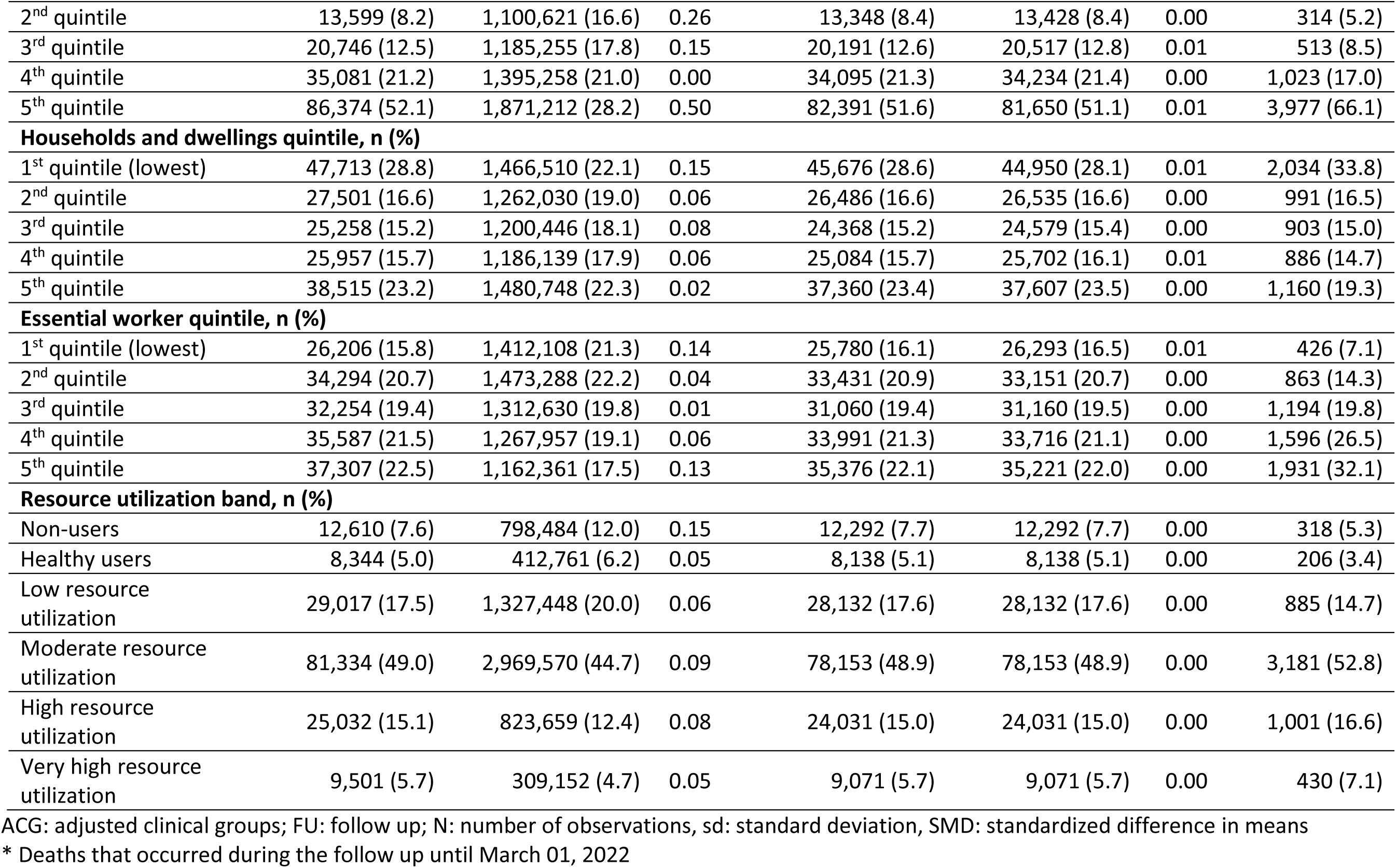
Demographics of individuals exposed to COVID-19 and unexposed individuals before and after matching.

| Characteristics | Pre-Matched |  |  | Matched |  |  | Unmatched |
| --- | --- | --- | --- | --- | --- | --- | --- |
|  | Exposed<br>N=165,838 | Unexposed<br>N=6,641,074 | SMD | Exposed<br>N=159,817 | Unexposed<br>N=159,817 | SMD | Exposed<br>N=6,021 |
| Age at index, years, mean (sd) | 40.4 ± 19.7 | 40.5 ± 22.8 | 0.01 | 40.4 ± 19.8 | 40.5 ± 19.9 | 0.01 | 39.9 ± 18.2 |
| Female sex, n (%) | 84,013 (50.7) | 3,362,838 (50.6) | 0.00 | 80,987 (50.7) | 80,987 (50.7) | 0.00 | 3,026 (50.3) |
| Rural, n (%) | 5,691 (3.4) | 95,577 (10.5) | 0.28 | 5,641 (3.5) | 5,655 (3.5) | 0.00 | 50 (0.8) |
| Immigrant, n (%) | 63,962 (38.6) | 1,167,364 (17.6) | 0.48 | 60,357 (37.8) | 59,435 (37.2) | 0.01 | 3,605 (59.9) |
| Frail, n (%) | 4,152 (2.5) | 124,329 (1.9) | 0.04 | 3,729 (2.3) | 5,580 (3.5) | 0.07 | 423 (7.0) |
| ACG, mean (sd) | 4.9 ± 3.5 | 4.1 ± 3.3 | 0.24 | 4.9 ± 3.5 | 4.7 ± 3.4 | 0.06 | 5.4 ± 3.4 |
| Died during follow-up*, n (%) | 3,357 (2.0) | 214,864 (3.2) | 0.08 | 3,222 (2.0) | 4,437 (2.8) | 0.05 | 135 (2.2) |
| **Neighbourhood income quintile, n (%)** |  |  |  |  |  |  |  |
| 1<sup>st</sup> quintile (lowest) | 40,878 (24.6) | 1,298,211 (19.5) | 0.12 | 39,182 (24.5) | 39,182 (24.5) | 0.00 | 1,696 (28.2) |
| 2<sup>nd</sup> quintile | 36,212 (21.8) | 1,303,219 (19.6) | 0.05 | 34,653 (21.7) | 34,653 (21.7) | 0.00 | 1,559 (25.9) |
| 3<sup>rd</sup> quintile | 36,205 (21.8) | 1,331,629 (20.1) | 0.04 | 34,731 (21.7) | 34,731 (21.7) | 0.00 | 1,474 (24.5) |
| 4<sup>th</sup> quintile | 29,252 (17.6) | 1,342,461 (20.2) | 0.07 | 28,385 (17.8) | 28,385 (17.8) | 0.00 | 867 (14.4) |
| 5<sup>th</sup> quintile (highest) | 23,291 (14.0) | 1,365,554 (20.6) | 0.17 | 22,866 (14.3) | 22,866 (14.3) | 0.00 | 425 (7.1) |
| **Age and labour force quintile, n (%)** |  |  |  |  |  |  |  |
| 1<sup>st</sup> quintile (lowest) | 59,199 (35.7) | 1,798,679 (27.1) | 0.19 | 57,196 (35.8) | 56,584 (35.4) | 0.01 | 2,017 (33.5) |
| 2<sup>nd</sup> quintile | 37,644 (22.7) | 1,351,752 (20.4) | 0.06 | 36,069 (22.6) | 36,258 (22.7) | 0.00 | 1,582 (26.3) |
| 3<sup>rd</sup> quintile | 26,766 (16.1) | 1,163,844 (17.5) | 0.04 | 25,800 (16.1) | 25,945 (16.2) | 0.00 | 954 (15.8) |
| 4<sup>th</sup> quintile | 22,439 (13.5) | 1,114,963 (16.8) | 0.09 | 21,664 (13.6) | 21,702 (13.6) | 0.00 | 754 (12.5) |
| 5<sup>th</sup> quintile (highest) | 18,896 (11.4) | 1,166,635 (17.6) | 0.18 | 18,245 (11.4) | 18,884 (11.8) | 0.01 | 667 (11.1) |
| **Material resource quintile, n (%)** |  |  |  |  |  |  |  |
| 1<sup>st</sup> quintile (lowest) | 27,993 (16.9) | 1,511,973 (22.8) | 0.15 | 27,408 (17.1) | 27,528 (17.2) | 0.00 | 594 (9.9) |
| 2<sup>nd</sup> quintile | 28,849 (17.4) | 1,390,533 (20.9) | 0.09 | 27,953 (17.5) | 28,033 (17.5) | 0.00 | 836 (13.9) |
| 3<sup>rd</sup> quintile | 32,509 (19.6) | 1,258,511 (19.0) | 0.02 | 31,358 (19.6) | 31,305 (19.6) | 0.00 | 1,215 (20.2) |
| 4<sup>th</sup> quintile | 34,440 (20.8) | 1,204,487 (18.1) | 0.07 | 32,887 (20.6) | 33,218 (20.8) | 0.01 | 1,558 (25.9) |
| 5<sup>th</sup> quintile | 41,153 (24.8) | 1,230,369 (18.5) | 0.15 | 39,368 (24.6) | 39,289 (24.6) | 0.00 | 1,771 (29.4) |
| **Racialized and newcomer populations quintile, n (%)** |  |  |  |  |  |  |  |
| 1<sup>st</sup> quintile (lowest) | 9,144 (5.5) | 1,043,527 (15.7) | 0.34 | 8,949 (5.6) | 9,544 (6.0) | 0.02 | 147 (2.4) |
| 2<sup>nd</sup> quintile | 13,599 (8.2) | 1,100,621 (16.6) | 0.26 | 13,348 (8.4) | 13,428 (8.4) | 0.00 | 314 (5.2) |
| 3<sup>rd</sup> quintile | 20,746 (12.5) | 1,185,255 (17.8) | 0.15 | 20,191 (12.6) | 20,517 (12.8) | 0.01 | 513 (8.5) |
| 4<sup>th</sup> quintile | 35,081 (21.2) | 1,395,258 (21.0) | 0.00 | 34,095 (21.3) | 34,234 (21.4) | 0.00 | 1,023 (17.0) |
| 5<sup>th</sup> quintile | 86,374 (52.1) | 1,871,212 (28.2) | 0.50 | 82,391 (51.6) | 81,650 (51.1) | 0.01 | 3,977 (66.1) |
| **Households and dwellings quintile, n (%)** |  |  |  |  |  |  |  |
| 1<sup>st</sup> quintile (lowest) | 47,713 (28.8) | 1,466,510 (22.1) | 0.15 | 45,676 (28.6) | 44,950 (28.1) | 0.01 | 2,034 (33.8) |
| 2<sup>nd</sup> quintile | 27,501 (16.6) | 1,262,030 (19.0) | 0.06 | 26,486 (16.6) | 26,535 (16.6) | 0.00 | 991 (16.5) |
| 3<sup>rd</sup> quintile | 25,258 (15.2) | 1,200,446 (18.1) | 0.08 | 24,368 (15.2) | 24,579 (15.4) | 0.00 | 903 (15.0) |
| 4<sup>th</sup> quintile | 25,957 (15.7) | 1,186,139 (17.9) | 0.06 | 25,084 (15.7) | 25,702 (16.1) | 0.01 | 886 (14.7) |
| 5<sup>th</sup> quintile | 38,515 (23.2) | 1,480,748 (22.3) | 0.02 | 37,360 (23.4) | 37,607 (23.5) | 0.00 | 1,160 (19.3) |
| **Essential worker quintile, n (%)** |  |  |  |  |  |  |  |
| 1<sup>st</sup> quintile (lowest) | 26,206 (15.8) | 1,412,108 (21.3) | 0.14 | 25,780 (16.1) | 26,293 (16.5) | 0.01 | 426 (7.1) |
| 2<sup>nd</sup> quintile | 34,294 (20.7) | 1,473,288 (22.2) | 0.04 | 33,431 (20.9) | 33,151 (20.7) | 0.00 | 863 (14.3) |
| 3<sup>rd</sup> quintile | 32,254 (19.4) | 1,312,630 (19.8) | 0.01 | 31,060 (19.4) | 31,160 (19.5) | 0.00 | 1,194 (19.8) |
| 4<sup>th</sup> quintile | 35,587 (21.5) | 1,267,957 (19.1) | 0.06 | 33,991 (21.3) | 33,716 (21.1) | 0.00 | 1,596 (26.5) |
| 5<sup>th</sup> quintile | 37,307 (22.5) | 1,162,361 (17.5) | 0.13 | 35,376 (22.1) | 35,221 (22.0) | 0.00 | 1,931 (32.1) |
| **Resource utilization band, n (%)** |  |  |  |  |  |  |  |
| Non-users | 12,610 (7.6) | 798,484 (12.0) | 0.15 | 12,292 (7.7) | 12,292 (7.7) | 0.00 | 318 (5.3) |
| Healthy users | 8,344 (5.0) | 412,761 (6.2) | 0.05 | 8,138 (5.1) | 8,138 (5.1) | 0.00 | 206 (3.4) |
| Low resource utilization | 29,017 (17.5) | 1,327,448 (20.0) | 0.06 | 28,132 (17.6) | 28,132 (17.6) | 0.00 | 885 (14.7) |
| Moderate resource utilization | 81,334 (49.0) | 2,969,570 (44.7) | 0.09 | 78,153 (48.9) | 78,153 (48.9) | 0.00 | 3,181 (52.8) |
| High resource utilization | 25,032 (15.1) | 823,659 (12.4) | 0.08 | 24,031 (15.0) | 24,031 (15.0) | 0.00 | 1,001 (16.6) |
| Very high resource utilization | 9,501 (5.7) | 309,152 (4.7) | 0.05 | 9,071 (5.7) | 9,071 (5.7) | 0.00 | 430 (7.1) |

ACG: adjusted clinical groups; FU: follow up; N: number of observations, sd: standard deviation, SMD: standardized difference in means
\* Deaths that occurred during the follow up until March 01, 2022

Most exposed individuals (96.0%) were matched to an unexposed individual. The matched cohort consisted of 319,634 individuals (159,817 exposed and 159,817 unexposed), with an average age of 40.4 ± 19.8 years, 50.7% were female, 24.5% lived in neighbourhoods with the lowest income levels, 20.7% had high or very high resource utilization, and 2.3% were frail (mean ACG score 4.9) (**Table 1**). The matched cohort was well-balanced, with no standardized differences above 0.1. Unmatched exposed individuals were more likely to live in lower income neighbourhoods, more likely to live in urban areas, had a higher mean ACG score, and were more likely to be frail at index date compared to matched exposed individuals (**Table 1**).

**Table S2** displays the demographic characteristics of the study cohort in the terminal phase. We identified 3,357 exposed individuals in the terminal phase and successfully matched 93.0% (N=3,114) of them to achieve balanced matches. The mean age of the matched cohort in the terminal phase was 76.9 ± 14.6 years, with 55.1% being female and 23.1% being immigrants. Unmatched individuals tended to be older, more likely to be frail, and more likely to be immigrants.

Within 14 days of the index date, 5.1% of the matched exposed cohort were hospitalized, and 26.5% of those were admitted to an intensive care unit (ICU). Overall, during the follow-up, 2% of the matched cohort died, including 20.1% of those who were hospitalized within 14 days of the index date without ICU admission and 39.1% of those admitted to an ICU.

### COVID-19-attributable healthcare costs

Mean (median) lengths of acute, post-acute, and terminal phases were 77 (79), 274 (280), and 35 (30) days for matched exposed. **Table 2** summarizes healthcare costs by phase of care. In the pre-index phase, the mean (95%CI) 10-day healthcare costs were similar for exposed and unexposed individuals ($107 ($103; $110) vs $106 ($103; $109) per person, respectively), resulting in $1 ($-4; $6) of COVID-19-attributable costs.

**Table 2.**
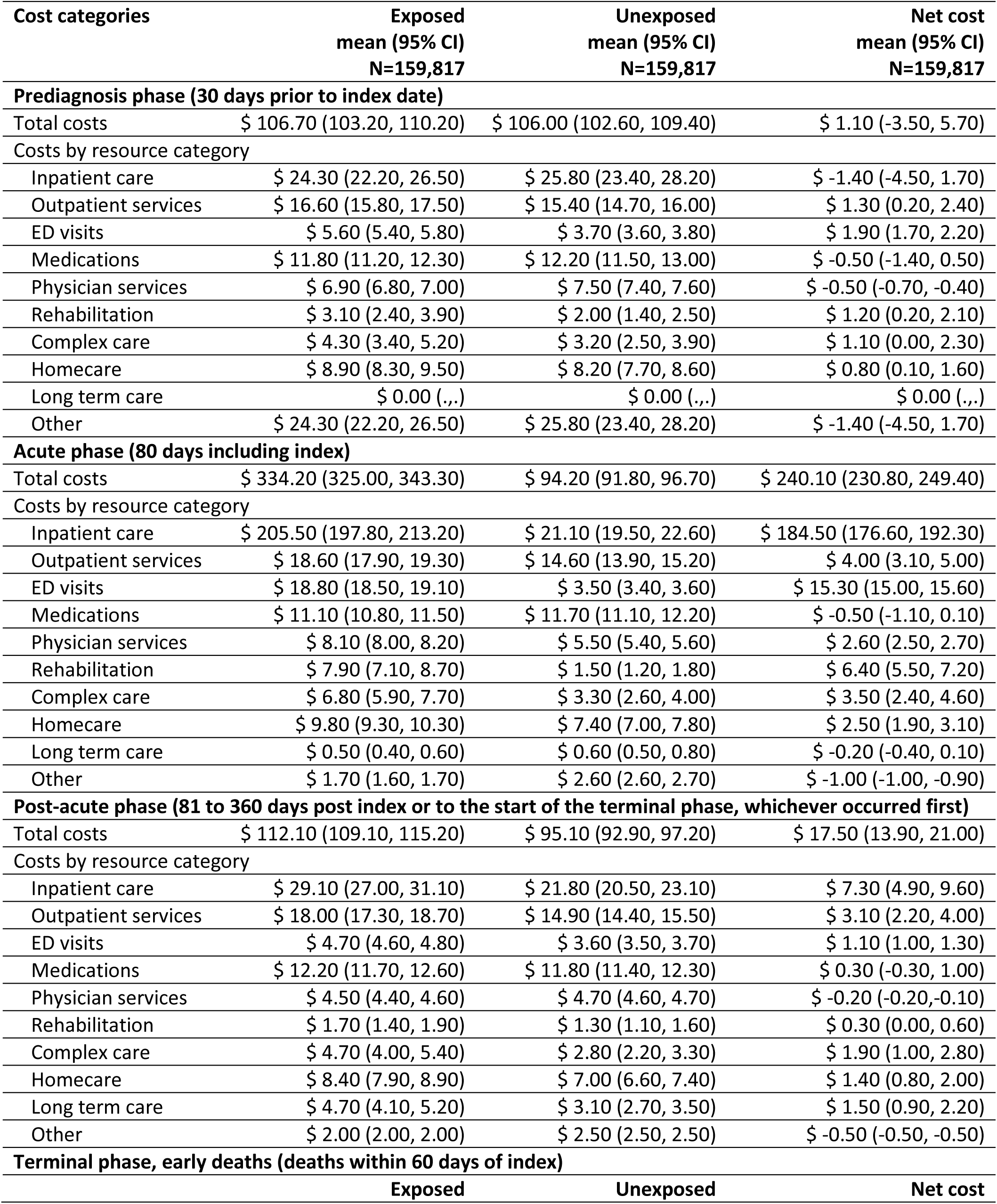
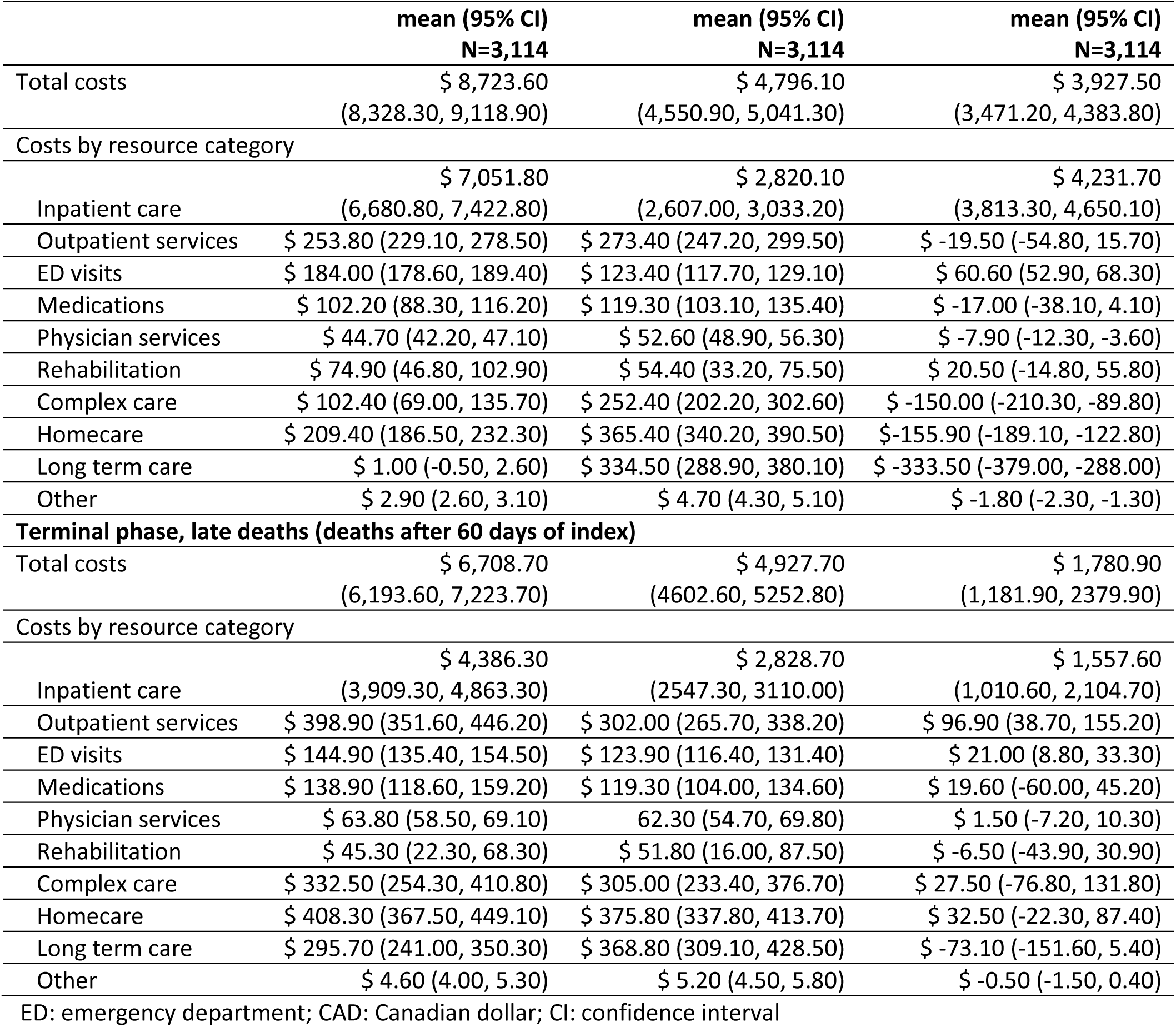
Total and COVID-19-attributable healthcare costs (2023 CAD) standardized to 10 days by phase of care.

During the acute phase, the mean 10-day health costs were $334 ($325; $343) per person for exposed and $94 ($92; $97) for unexposed individuals, resulting in $240 ($231; $249) of COVID-19-attributable costs. Most acute phase costs were due to inpatient care (77%), followed by emergency department visits (6%) and rehabilitation services (3%) (**Figure 2).**

**Figure 2.**
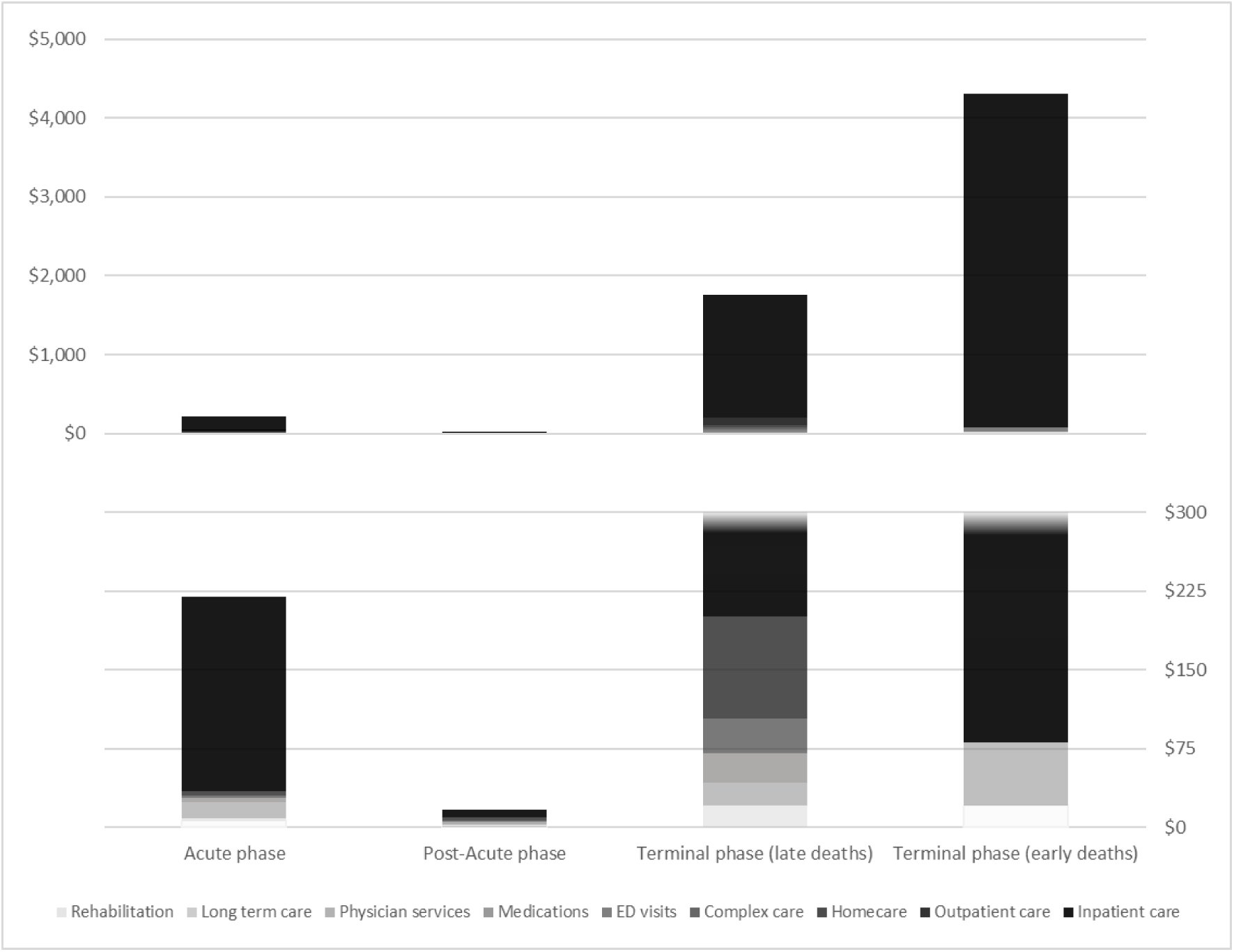
Source of COVID-19-attributable healthcare costs (2023 CAD) standardized to 10 days by phases-of-care. CAD: Canadian dollar; ED: emergency department Figure displays cost categories by phase of care. The Y-axis (bottom panel) had been truncated at $300 to improve visualization of the post-acute phase costs. The upper panel shows the full-length Y-axis without truncation.

During the post-acute phase, the mean 10-day health costs were $112 ($109; $115) per person for exposed and $95 ($93; $97) for unexposed individuals, resulting in $18 ($14; $21) of COVID-19-attributable costs. Most post-acute phase costs were due to inpatient care (42%), followed by outpatient services (18%) and complex care (11%) (**Figure 2**).

During the terminal phase, among individuals with early deaths, the mean 10-day health costs were $8,724 ($8,328; $9,119) per person for exposed and $4,796 ($4,551; $5,041) for unexposed individuals, resulting in $3,928 ($3,471; $4,384) of COVID-19-attributable costs. Among individuals with late deaths, the mean 10-day health costs were $6,709 ($6,194; $7,224) per person for exposed and $4,928 ($4,603; $5,253) for unexposed individuals, resulting in $1,781 ($1,182; $2,380) of COVID-19-attributable costs. Most terminal phase costs were due to inpatient care for both early (100%) and late deaths (>87%) (**Figure 2**).

Mean (95%CI) attributable 10-day COVID-19 costs were lower for females than males for the acute care phase at $193 ($182, $204) vs. $289 ($274, $304), but higher for females than males in the post-acute care phase $21 ($16, $26) vs. $14 ($9, $19). Total mean attributable costs in both acute and post-acute care phases were higher among older age groups, among those in the lower income quintiles, and in the two highest resource utilization bands (**Tables S3-S6**).

Healthcare costs were not sensitive to variations in the pre-index phase length. However, compared to the phase lengths in the main analysis, costs doubled for the acute and post-acute phases, when we considered shorter duration (30 days) for acute phase, with inpatient care remaining the main cost driver (**Table S7**). This occurred because costs initially captured under the acute phase were now shifted to the post-acute phase (**Figure 3**). Finally, mean cumulative costs adjusted for survival at 360 days were $2,553 ($2,348; $2,756) with lower costs for females at $2,194 ($1,945; $2,446) vs. males at $2,921 ($2,602; $3,241**) (Table 3).**

**Figure 3.**
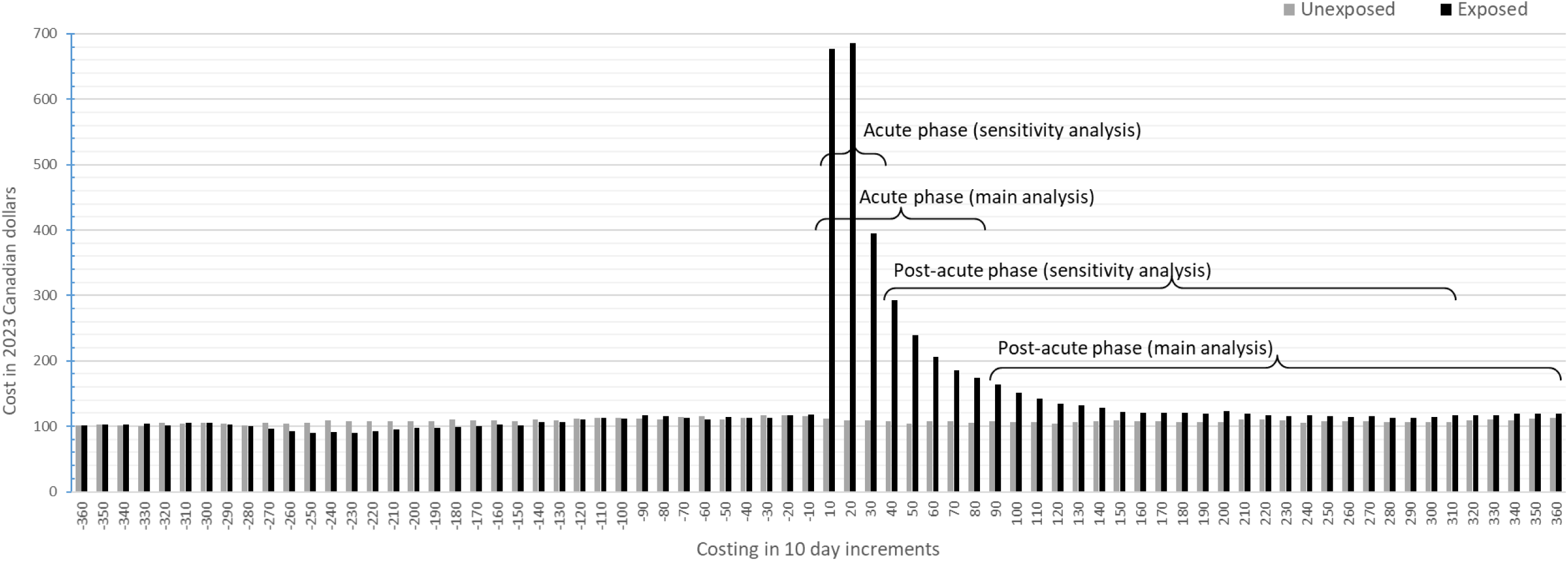
Healthcare costs (2023 CAD) per 10-day intervals for the matched exposed and unexposed individuals. Note: Costs of individuals who died during the follow up were removed 60 days prior to death consistent with the terminal phase length for late deaths.

**Table 3.** COVID-19-attributable costs (2023 CAD), adjusted for survival, total and stratified by age and sex.

|  | **30 days**<br>**Mean (95% CI)** | **90 days**<br>**Mean (95% CI)** | **180 days**<br>**Mean (95% CI)** | **360 days**<br>**Mean (95% CI)** |
| --- | --- | --- | --- | --- |
| **Total** | \$ 790 (753, 826) | \$ 2,043 (1,949, 2,142) | \$ 2,215 (2,083, 2,346) | \$ 2,553 (2,348, 2,756) |
| **Age** |  |  |  |  |
| <2 years* | \$ 404 (283, 526) | \$ 1,094 (756, 1,432) | \$ 1,233 (766, 1,701) | \$1,512 (786, 2,240) |
| 2-4 years* | \$ 59 (20, \$97) | \$168 (47, 290) | \$ 273 (-23, 570) | \$484 (-161, 1,132) |
| 5-11 years* | \$ 69 (35, 104) | \$ 185 (89, 281) | \$ 189 (55, 323) | \$ 196 (-14, 406) |
| 12-17 years* | \$ 45 (21, 69) | \$ 122 (52, 193) | \$ 143 (18, 269) | \$ 184 (-50, 420) |
| 18-29 years | \$ 89 (72, 106) | \$ 239 (190, 288) | \$ 259 (179, 337) | \$ 293 (134, 449) |
| 30-49 years | \$ 348 (311, 385) | \$ 932 (832, 1015) | \$ 1,030 (888, 1172) | \$ 1,224 (1,000, 1,449) |
| 50-69 years | \$ 1,366 (1,27, 1,461) | \$ 3,556 (3,305, 3656) | \$ 3,789 (3,454, 4125) | \$ 4,247 (3,748, 4,745) |
| 70+ years | \$ 1,672 (1,532, 1,813) | \$ 3,958 (3,633, 3,841) | \$ 4,242 (3,804, 4,680) | \$13,841 (12,193, 15,489) |
| **Sex** |  |  |  |  |
| Females | \$ 613 (572, 655) | \$ 1,611 (1,504, 1,753) | \$ 1,807 (1,651, 1,963) | \$ 2,194 (1,945, 2,446) |
| Males | \$ 971 (911, 1,031) | \$ 2,490 (2,335, 2,603) | \$ 2,637 (2,426, 2,848) | \$ 2,921 (2,602, 3,241) |

CAD: Canadian dollar; CI: confidence interval
Costs were rounded to the nearest dollar
\*calculated as if no one died in these categories due to small cells/ unable to model a stable attributable cost estimate for the terminal phases

Healthcare costs of individuals who were hospitalized within 14 days of the index date with or without ICU admission are summarized in **Table S8**.

## Discussion

We sought to characterize the acute and long-term healthcare costs attributable to COVID-19 in Ontario, Canada, among individuals who tested positive for SARS-CoV-2, compared to a group of individuals who were unexposed to SARS-CoV-2.

Exposed individuals had higher costs than unexposed individuals post index date, with the largest difference occurring during the acute phase. While the cost differential (attributable cost) was reduced in the post-acute phase, mean costs remained greater among exposed individuals compared to unexposed individuals until the end of follow-up at 1 year post index date.

The relative contributions of specific cost categories to COVID-19-attributable costs varied by phase. The leading cost category in COVID-19-attributable cost across all phases was inpatient care, contributing 77% in the acute phase and 42% during the post-acute phase. This is consistent with descriptions of the natural history of SARS-CoV-2 acute infections and subsequent PCC. Acute infection often requires care to address acute symptoms, including potential respiratory distress, that would be best addressed in hospital depending on severity, while the PCC consists of several linked syndromic conditions that are more likely to require complex care, rehabilitation, and outpatient visits [36].

We found the duration of acute illness to be longer than reported in other studies. Using joinpoint regression analysis, we determined the acute phase length to be 80 days. This is longer than the 30 days used to define the onset of a post-acute period by Sagy et al.[14], but lends further empirical support to a growing global consensus that PCC is defined by symptoms that persist more than 12 weeks after infection onset [37–39].

The mean cumulative cost at 360 days for COVID-19 was $2,553 per person tested positive for SARS-CoV-2, which translates to $408M for the entire exposed cohort of 159,817 individuals. Meanwhile, the estimated cost for PCC ($18 attributable cost in the post-acute phase per person, per 10 days), would amount to about $75M for the entire exposed cohort over a year.

Our findings are comparable to those of a previous COVID-19 costing study in Canada. Tsui et al. estimated healthcare costs associated with COVID-19 during the first wave of the pandemic in two Canadian provinces, British Columbia and Ontario [40]. The authors stratified costs based on the level of initial care required, including community, long-term care, hospital, and ICU settings. In Ontario, the net costs (in 2020 CAD) during the first 30 days of diagnosis were $15,750 for individuals who were hospitalized and $56,088 for those admitted to the ICU. The cumulative 120-day costs were $28,329 and $96,308, respectively. In our study, after stratifying by the level of initial care received (**Table S8**), the net cost over 360-day period were $30,147 and $105,677 for individuals who were hospitalized and admitted to ICU, respectively. The higher costs in our study, due to the longer time horizon, suggest the ongoing impact of PCC, particularly among those with initially severe disease. Further, we found that even after matching for comorbidities, frailty, and other factors, healthcare costs remained higher for lower-income quintiles. This finding aligns with a study conducted in Ontario, showing that lower-income individuals have a higher mortality risk, a factor that likely contributes to their increased end-of-life healthcare costs [41].

Our study has several limitations. Consistent with the health system perspective, outcome data are limited to OHIP administrative data, and therefore do not include healthcare-related costs not covered by Ontario’s public insurance system, such as private caregivers, medications for most individuals under 65 years, copayments for medications, costs covered by private insurance, or community-level services such as supportive housing. Also, while the vast majority (96%) of exposed individuals were matched to unexposed individuals, unmatched exposed individuals were more likely to live in lower income neighbourhoods, more likely to live in urban areas, had higher mean ACG score and were more likely to be frail at index date compared to the matched exposed, making findings less generalizable to individuals with these characteristics. Finally, our study reflects early pandemic experiences, when the likelihood of severe illness requiring hospitalization was higher, leading to increased healthcare costs. COVID-19 and associated healthcare costs may differ in contemporary times with the emergence of novel variants, vaccinations, treatments, and changes in clinical practice which were not assessed in this study.

Our study has important strengths. First, because Ontario has a single public payer system, most of the community dwelling population was eligible for this study. The cohort was well matched across various clinical and demographic variables. Notably, the absence of COVID-19-attributable costs at baseline suggested minimal residual confounding. Furthermore, matching to historic controls ensured that changes in healthcare delivery during the pandemic did not affect the unexposed group, which could bias attributable cost estimates for COVID-19. Second, the use of several linked administrative databases allowed for the characterization of COVID-19-attributable costs across a broad range of cost categories. Our analysis goes beyond inpatient costs, providing a comprehensive view of COVID-19 healthcare costs. Finally, a phase-of-care approach enabled a comparison of acute versus post-acute costs and the use of joinpoint regression allowed for a data-driven approach to defining phase length.

Our findings offer insights into clinically relevant phase-specific costs, which can inform budget planning to ensure each healthcare sector is appropriately resourced. Inpatient care emerges as a major cost category, emphasizing the need for hospital resources. Policymakers can use these findings to prioritize investments as well as to assess the value of COVID-19 interventions to support current policy decision-making and pandemic planning.

## Conclusion

Our study found that SARS-CoV-2 infection is associated with increased healthcare costs in the year following onset, with differential cost patterns in the acute and post-acute phases, consistent with the evolving clinical understanding of the PCC. Our findings have important implications for stakeholders responding to the PCC at the health system level.

## Author Contributions

*Conceptualization:* J Kwong, S Mishra, J Naveed, S Hind; B Sander;

*Design and Methodology:* J Kwong, S Mishra, K Quinn, B Sander; *Analysis:* S Swayze;

*Writing – Original Draft:* R Duchen, Y Sahakyan;

*Writing – Review & Editing and Interpretation and final approval:* All authors;

*Supervision*: J Kwong, S Mishra, K Quinn, B Sander;

*Funding acquisition:* J Kwong, S Mishra, B Sander.

## Acknowledgements

This document used data adapted from the Statistics Canada 2016 Census Area Profiles and the Statistics Canada Postal Code^OM^ Conversion File, which is based on data licensed from Canada Post Corporation, and/or data adapted from the Ontario Ministry of Health Postal Code Conversion File, which contains data copied under license from ©Canada Post Corporation and Statistics Canada.

Parts of this material are based on data and/or information compiled and provided by Statistics Canada, the Canadian Institute for Health Information (CIHI), Ontario Health (OH), and the Ontario Ministry of Health as well as Immigration, Refugees and Citizenship Canada (IRCC) current to September 2020. The analyses, conclusions, opinions, and statements expressed herein are solely those of the authors and do not reflect those of the funding or data sources, no endorsement is intended or should be inferred.

We thank the Toronto Community Health Profiles Partnership for providing access to the Ontario Marginalization Index.

The authors thank Poornima Goudar and Marian Hassan for helping with editing the manuscript and Hong Lu for her coding contributions during the analysis.

## Data availability

The dataset from this study is held securely in a coded form at ICES. While legal data-sharing agreements between ICES and data providers (e.g., healthcare organizations and government) prohibit ICES from making the dataset publicly available, access may be granted to those who meet pre-specified criteria for confidential access, available at www.ices.on.ca/DAS. The full dataset creation plan and underlying analytic code are available from the authors upon request, understanding that computer programs may rely upon coding templates or macros that are unique to ICES and are therefore either inaccessible or may require modification.

## Ethics approval

This study was approved by the University Health Network Research Ethics Board. ICES is a prescribed entity under Ontario’s Personal Health Information Protection Act (PHIPA). Section 45 of the PHIPA authorizes the ICES to collect personal health information, without consent, for the purpose of analysis or compiling statistical information with respect to the management of, evaluation or monitoring of the allocation of resources to, or planning for all or part of the health system. Projects that use data collected by ICES under section 45 of PHIPA and use no other data are exempt from the REB review. The use of the data in this project is authorized under Section 45 and approved by the ICES Privacy and Legal Office.

## Supplementary Materials

**Table S1.** ICES Datasets used in this study.

| List of datasets | Full name | Description |
| --- | --- | --- |
| C19INTGR | COVID19 Integrated Testing Data | Used to determine those with a positive C19 infection |
| CCM | Case and Contact Management System | Used to determine those with a positive C19 infection |
| CCRS | Continuing Care Reporting System | Used to determine those living in Long-Term care facilities and part of the healthcare cost calculation |
| CENSUS | Ontario Census Area Profiles | used to create the Essential worker quintile |
| DAD | Discharge Abstract Database DAD | Part of the health care cost calculations and hospitalizations |
| HCD | Home Care Database | Part of the health care cost calculations |
| CIC/IRCC | IRCC Permanent Residents database | Used to determine immigration status |
| NACRS | National Ambulatory Care Reporting System | Part of the health care cost calculations |
| ODB | Ontario Drug Benefit Claims | Part of the health care cost calculations |
| OHIP | Ontario Health Insurance Plan Claims Database | Part of the health care cost calculations |
| OLISC19 | OLIS COVID-19 Laboratory Data | Used to determine those with a positive C19 infection |
| OMHRS | Ontario Mental Health Reporting System | Part of the health care cost calculations |
| ON-MARG | Ontario Marginalization Index | Used to create the Age and labour force quintiles, Material resource quintiles, Racialized and newcomer populations quintile, and Households and dwellings quintile |
| PCCF | Postal Code Conversion File | Used to determine postal code for Ontario residency |
| RPDB | Registered Persons Database | Demographics information on birth date, sex, ohip eligibility, date of last contact with the healthcare system, and death date |
| SDS | Same Day Surgery Database (Annual) | Part of the health care cost calculations |
| ACG | ACG macro | Used in combination with OHIP, DAD, and NACRS to calculate ACGs, RUB, and frailty |
| NRS | National Rehabilitation Reporting System | Part of the health care cost calculations |
| NDFP | New Drug Funding Program | Part of the health care cost calculations |
| ADP | Assistive Devices Program | Part of the health care cost calculations |
| INST | Facilities | Used to determine those living in Long-Term care facilities |

**Source:** For more information please check https://datadictionary.ices.on.ca/Applications/DataDictionary/Default.aspx

**Table S2.** Demographics of individuals exposed to COVID-19 and unexposed individuals in the terminal phase.

| Characteristics | Matched |  | SMD | Unmatched |
| --- | --- | --- | --- | --- |
|  | Exposed<br>N=3,114 | Unexposed<br>N=3,114 |  | Exposed<br>N=243 |
| Age at index, years, mean (sd) | 76.9 ± 14.6 | 75.4 ± 16.1 | 0.10 | 81.7 ± 16.6 |
| Female sex, n (%) | 1,399 (44.9) | 1,399 (44.9) | 0.00 | 109 (44.9) |
| Rural, n (%) | 116 (3.7) | 104 (3.3) | 0.02 | <=5 (...) |
| Immigrant, n (%) | 720 (23.1) | 720 (23.1) | 0.00 | 83 (34.2) |
| Frail, n (%) | 1,056 (33.9) | 1,136 (36.5) | 0.05 | 136 (56.0) |
| ACG, median (IQR) | 9.0 (6.0-12.0) | 9.0 (6.0-12.0) | 0.09 | 9.0 (6.0-12.0) |
| **Neighbourhood income quintile, n (%)** |  |  |  |  |
| 1<sup>st</sup> quintile (lowest) | 889 (28.5) | 889 (28.5) | 0.00 | 68 (28.0) |
| 2<sup>nd</sup> quintile | 738 (23.7) | 738 (23.7) | 0.00 | 39 (16.0) |
| 3<sup>rd</sup> quintile | 631 (20.3) | 631 (20.3) | 0.00 | 78 (32.1) |
| 4<sup>th</sup> quintile | 435 (14.0) | 435 (14.0) | 0.00 | 22 (9.1) |
| 5<sup>th</sup> quintile (highest) | 421 (13.5) | 421 (13.5) | 0.00 | 36 (14.8) |
| **Age and labour force quintile, n (%)** |  |  |  |  |
| 1<sup>st</sup> quintile (lowest) | 619 (19.9) | 602 (19.3) | 0.01 | 58 (23.9) |
| 2<sup>nd</sup> quintile | 577 (18.5) | 584 (18.8) | 0.01 | 49 (20.2) |
| 3<sup>rd</sup> quintile | 542 (17.4) | 550 (17.7) | 0.01 | 22 (9.1) |
| 4<sup>th</sup> quintile | 533 (17.1) | 559 (18.0) | 0.02 | 73 (30.0) |
| 5<sup>th</sup> quintile (highest) | 823 (26.4) | 803 (25.8) | 0.01 | 35 (14.4) |
| **Material resource quintile, n (%)** |  |  |  |  |
| 1<sup>st</sup> quintile (lowest) | 529 (17.0) | 550 (17.7) | 0.02 | 44 (18.1) |
| 2<sup>nd</sup> quintile | 513 (16.5) | 503 (16.2) | 0.01 | 44 (18.1) |
| 3<sup>rd</sup> quintile | 565 (18.1) | 574 (18.4) | 0.01 | 62 (25.5) |
| 4<sup>th</sup> quintile | 654 (21.0) | 633 (20.3) | 0.02 | 38 (15.6) |
| 5<sup>th</sup> quintile | 833 (26.8) | 838 (26.9) | 0.00 | 49 (20.2) |
| **Racialized and newcomer populations quintile, n (%)** |  |  |  |  |
| 1<sup>st</sup> quintile (lowest) | 267 (8.6) | 242 (7.8) | 0.03 | 19 (7.8) |
| 2<sup>nd</sup> quintile | 371 (11.9) | 339 (10.9) | 0.03 | 40 (16.5) |
| 3<sup>rd</sup> quintile | 488 (15.7) | 495 (15.9) | 0.01 | 31 (12.8) |
| 4<sup>th</sup> quintile | 680 (21.8) | 693 (22.3) | 0.01 | 47 (19.3) |
| 5<sup>th</sup> quintile | 1,288 (41.4) | 1,329 (42.7) | 0.03 | 100 (41.2) |
| **Households and dwellings quintile, n (%)** |  |  |  |  |
| 1<sup>st</sup> quintile (lowest) | 541 (17.4) | 549 (17.6) | 0.01 | 51 (21.0) |
| 2<sup>nd</sup> quintile | 442 (14.2) | 442 (14.2) | 0.00 | 22 (9.1) |
| 3<sup>rd</sup> quintile | 491 (15.8) | 478 (15.4) | 0.01 | 33 (13.6) |
| 4<sup>th</sup> quintile | 557 (17.9) | 528 (17.0) | 0.02 | 46 (18.9) |
| 5<sup>th</sup> quintile | 1,063 (34.1) | 1,101 (35.4) | 0.03 | 85 (35.0) |
| **Essential worker quintile, n (%)** |  |  |  |  |
| 1<sup>st</sup> quintile (lowest) | 608 (19.5) | 616 (19.8) | 0.01 | 38 (15.6) |
| 2<sup>nd</sup> quintile | 601 (19.3) | 599 (19.2) | 0.00 | 67 (27.6) |
| 3<sup>rd</sup> quintile | 612 (19.7) | 594 (19.1) | 0.01 | 30 (12.3) |
| 4<sup>th</sup> quintile | 654 (21.0) | 654 (21.0) | 0.00 | 55 (22.6) |
| 5<sup>th</sup> quintile | 631 (20.3) | 636 (20.4) | 0.00 | 49 (20.2) |
| **Resource utilization band, n (%)** |  |  |  |  |
| Non-users | 35 (1.1) | 35 (1.1) | 0.00 | 12 (4.9)* |
| Healthy Users | 10 (0.3) | 10 (0.3) | 0.00 |  |
| Low resource utilization | 79 (2.5) | 79 (2.5) | 0.00 | 12 (4.9) |
| Moderate resource utilization | 832 (26.7) | 832 (26.7) | 0.00 | 54 (22.2) |
| High resource utilization | 658 (21.1) | 658 (21.1) | 0.00 | 52 (21.4) |
| Very high resource utilization | 1,500 (48.2) | 1,500 (48.2) | 0.00 | 113 (46.5) |

ACG: adjusted clinical groups; IQR; interquartile range; N: number of observations, sd: standard deviation, SMD: standardized difference in means
\*two categories were combined because of small number

**Table S3.** Total and COVID-19-attributable healthcare costs (2023 CAD) standardized to 10 days stratified by age.

| Age categories | N | Exposed<br>mean (95% CI) | N | Unexposed<br>mean (95% CI) | Net cost<br>mean (95% CI) |
| --- | --- | --- | --- | --- | --- |
| **Prediagnosis phase (30 days prior to index date)** |  |  |  |  |  |
| 0 to <2 years old* | 1,138 | \$ 100.90 (70.10, 131.70) | 1,139 | \$ 163.30 (95.40, 231.20) | \$ -62.40 (-137.10, 12.20) |
| 2-4 years old* | 2,152 | \$ 27.00 (16.80, 37.20) | 2,152 | \$ 39.90 (28.50, 51.30) | \$ -13.00 (-27.90, 2.00) |
| 5-11 years old* | 6,675 | \$ 23.70 (17.90, 29.50) | 6,675 | \$ 23.30 (20.30, 26.20) | \$ 0.40 (-5.90, 6.70) |
| 12-17 years old* | 8,519 | \$ 32.00 (26.50, 37.50) | 8,519 | \$ 34.00 (28.30, 39.60) | \$ -2.00 (-9.40, 5.50) |
| 18-29 years old | 35,515 | \$ 40.20 (36.80, 43.70) | 35,517 | \$ 44.90 (41.40, 48.30) | \$ -4.60 (-9.20, 0.00) |
| 30-49 years old | 51,773 | \$ 64.70 (60.90, 68.50) | 51,809 | \$ 68.70 (64.90, 72.50) | \$ -4.00 (-9.30, 1.20) |
| 50-69 years old | 41,601 | \$ 132.20 (124.4, 140.10) | 41,903 | \$ 133.30 (125.90, 140.80) | \$ -1.00 (-11.50, 9.40) |
| 70+ years old | 10,820 | \$ 553.90 (520.80, 586.90) | 12,026 | \$ 455.20 (424.70, 485.60) | \$ 100.40 (57.90, 143.00) |
| **Acute phase (80 days including index)** |  |  |  |  |  |
| 0 to <2 years old* | 1,138 | \$ 189.20 (150.40, 227.90) | 1,139 | \$ 54.40 (41.70, 67.10) | \$ 134.80 (94.40, 175.30) |
| 2-4 years old* | 2,152 | \$ 49.50 (37.40, 61.50) | 2,152 | \$ 29.90 (24.10, 35.80) | \$ 19.50 (6.80, 32.30) |
| 5-11 years old* | 6,675 | \$ 45.00 (33.80, 56.20) | 6,675 | \$ 21.90 (19.40, 24.50) | \$ 23.10 (11.60, 34.60) |
| 12-17 years old* | 8,519 | \$ 47.70 (40.8, 54.50) | 8,519 | \$ 32.60 (28.30, 36.90) | \$ 15.00 (7.00, 23.10) |
| 18-29 years old | 35,512 | \$ 70.60 (65.50, 75.60) | 35,517 | \$ 41.00 (38.20, 43.70) | \$ 29.60 (23.90, 35.30) |
| 30-49 years old | 51,762 | \$ 172.70 (162.50, 183.00) | 51,799 | \$ 59.70 (56.90, 62.50) | \$ 113.00 (102.40, 123.60) |
| 50-69 years old | 41,484 | \$ 534.90 (509.90, 559.90) | 41,887 | \$ 120.50 (115.00, 126.00) | \$ 414.40 (389.00, 439.80) |
| 70+ years old | 10,563 | \$ 1,710.60 (1,638.50, 1,782.70) | 11,969 | \$ 408.90 (388.10, 429.70) | \$ 1,302.30 (1,228.50, 1,376.10) |
| **Post-acute phase (81 to 360 days post index or to the start of the terminal phase, whichever occurred first)** |  |  |  |  |  |
| 0 to <2 years old* | 1,138 | \$ 49.20 (35.10, 63.20) | 1,139 | \$ 33.70 (29.90, 37.50) | \$ 15.50 (1.10, 29.90) |
| 2-4 years old* | 2,152 | \$ 39.30 (19.60, 59.10) | 2,152 | \$ 27.60 (23.00, 32.20) | \$ 11.70 (-7.70, 31.20) |
| 5-11 years old* | 6,675 | \$ 21.40 (17.80, 24.90) | 6,675 | \$ 21.00 (18.80, 23.20) | \$ 0.40 (-3.80, 4.60) |
| 12-17 years old* | 8,519 | \$ 33.40 (28.10, 38.60) | 8,519 | \$ 31.00 (27.80, 34.30) | \$ 2.30 (-3.80, 8.40) |
| 18-29 years old | 35,507 | \$ 43.40 (40.90, 45.90) | 35,514 | \$ 41.20 (38.90, 43.50) | \$ 2.20 (-1.20, 5.50) |
| 30-49 years old | 51,751 | \$ 70.20 (66.60, 73.80) | 51,791 | \$ 59.70 (57.40, 61.90) | \$ 10.50 (6.30, 14.70) |
| 50-69 years old | 41,421 | \$ 146.40 (139.40, 153.40) | 41,841 | \$ 122.70 (118.00, 127.30) | \$ 23.70 (15.60, 31.80) |
| 70+ years old | 10,373 | \$ 565.10 (536.90, 593.30) | 11,806 | \$ 421.00 (401.60, 440.30) | \$ 145.70 (112.70, 178.70) |
| **Terminal phase, early deaths (deaths within 60 days of index)\*** |  |  |  |  |  |
| 18-29 years old | 8 | \$ 7,600.50 (130.30, 15,070.80) | 29 | \$ 4,299.10 (1,223.30, 7,374.90) | \$ -1,387.10 (-8,157.30, 5,383.00) |
| 30-49 years old | 52 | \$ 10,7350 (7,478.10, 13,991.80) | 106 | \$ 4,217.90 (2,880.80, 5,555.00) | \$ 6,506.60 (3,089.80, 9,923.40) |
| 50-69 years old | 426 | \$ 13,465.30 (12,415.00, 14,515.50) | 430 | \$ 5,244.40 (4,721.50, 5,767.40) | \$ 8,219.80 (7,045.80, 9,393.70) |
| 70+ years old | 1,432 | \$ 7,251.60 (6,870.40, 7,632.70) | 1,348 | \$ 4,689.00 (4,411.50, 4,966.40) | \$ 2,541.40 (2,089.20, 2,993.60) |
| **Terminal phase, late deaths (deaths after 60 days of index)\*** |  |  |  |  |  |
| 18-29 years old | 21 | \$ 4,430.30 (1,272.10, 7,588.60) | 22 | \$ 6,667.60 (1,846.70, 11,488.40) | \$ -1,720.40 (-6,876.40, 3,435.60) |
| 30-49 years old | 56 | \$ 7,765.40 (4,569.20, 10,961.70) | 60 | \$ 4,758.60 (3,430.90, 6,086.30) | \$ 2,991.30 (-433.60, 6,416.30) |
| 50-69 years old | 278 | \$ 8,612.10 (7,302.50, 9,921.80) | 257 | \$ 5,710.30 (4,789.40, 6,631.10) | \$ 2,915.00 (1,355.10, 4,474.90) |
| 70+ years old | 838 | \$ 6,070.60 (5,526.40, 6,614.80) | 851 | \$ 4,611.60 (4,288.10, 4,935.10) | \$ 1,451.40 (830.20, 2,072.60) |

CI: confidence interval
\* unable to model a stable attributable cost estimate for age <18 years due to small cells for terminal phases

**Table S4.**
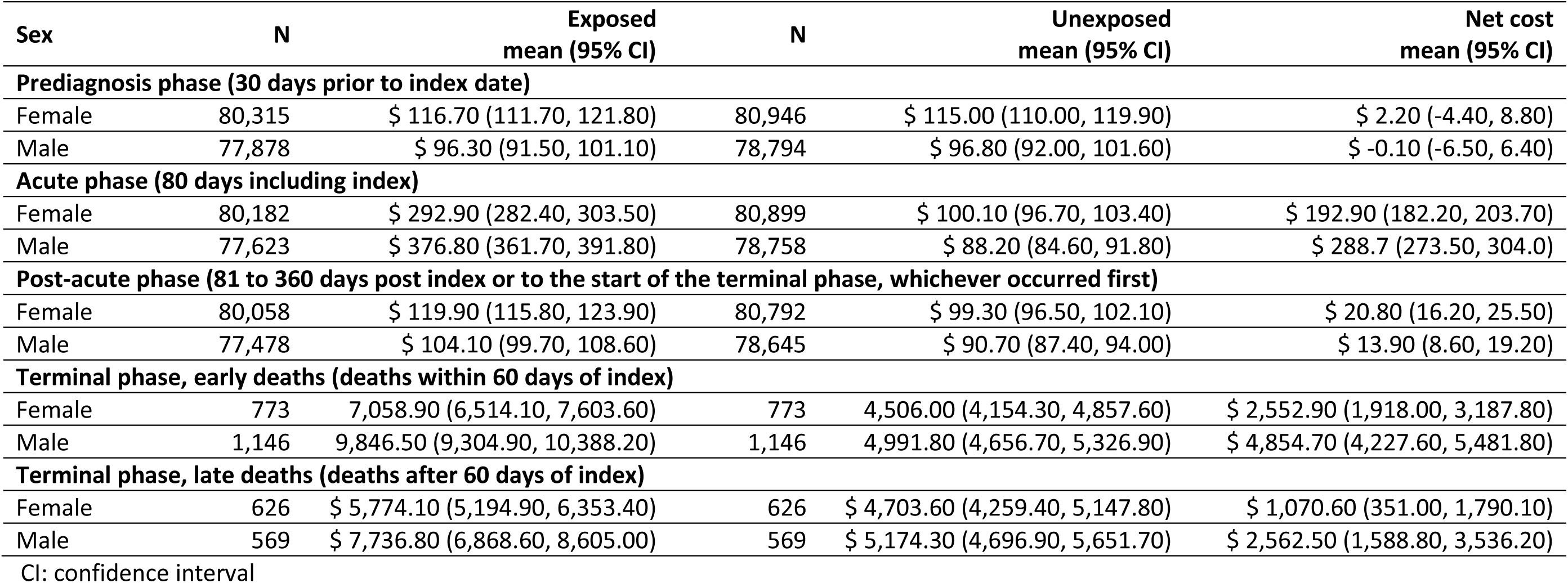
Total and COVID-19-attributable healthcare costs (2023 CAD) standardized to 10 days stratified by sex.

| Sex | N | Exposed<br>mean (95% CI) | N | Unexposed<br>mean (95% CI) | Net cost<br>mean (95% CI) |
| --- | --- | --- | --- | --- | --- |
| **Prediagnosis phase (30 days prior to index date)** |  |  |  |  |  |
| Female | 80,315 | \$ 116.70 (111.70, 121.80) | 80,946 | \$ 115.00 (110.00, 119.90) | \$ 2.20 (-4.40, 8.80) |
| Male | 77,878 | \$ 96.30 (91.50, 101.10) | 78,794 | \$ 96.80 (92.00, 101.60) | \$ -0.10 (-6.50, 6.40) |
| **Acute phase (80 days including index)** |  |  |  |  |  |
| Female | 80,182 | \$ 292.90 (282.40, 303.50) | 80,899 | \$ 100.10 (96.70, 103.40) | \$ 192.90 (182.20, 203.70) |
| Male | 77,623 | \$ 376.80 (361.70, 391.80) | 78,758 | \$ 88.20 (84.60, 91.80) | \$ 288.7 (273.50, 304.0) |
| **Post-acute phase (81 to 360 days post index or to the start of the terminal phase, whichever occurred first)** |  |  |  |  |  |
| Female | 80,058 | \$ 119.90 (115.80, 123.90) | 80,792 | \$ 99.30 (96.50, 102.10) | \$ 20.80 (16.20, 25.50) |
| Male | 77,478 | \$ 104.10 (99.70, 108.60) | 78,645 | \$ 90.70 (87.40, 94.00) | \$ 13.90 (8.60, 19.20) |
| **Terminal phase, early deaths (deaths within 60 days of index)** |  |  |  |  |  |
| Female | 773 | 7,058.90 (6,514.10, 7,603.60) | 773 | 4,506.00 (4,154.30, 4,857.60) | \$ 2,552.90 (1,918.00, 3,187.80) |
| Male | 1,146 | 9,846.50 (9,304.90, 10,388.20) | 1,146 | 4,991.80 (4,656.70, 5,326.90) | \$ 4,854.70 (4,227.60, 5,481.80) |
| **Terminal phase, late deaths (deaths after 60 days of index)** |  |  |  |  |  |
| Female | 626 | \$ 5,774.10 (5,194.90, 6,353.40) | 626 | \$ 4,703.60 (4,259.40, 5,147.80) | \$ 1,070.60 (351.00, 1,790.10) |
| Male | 569 | \$ 7,736.80 (6,868.60, 8,605.00) | 569 | \$ 5,174.30 (4,696.90, 5,651.70) | \$ 2,562.50 (1,588.80, 3,536.20) |

CI: confidence interval

**Table S5.**
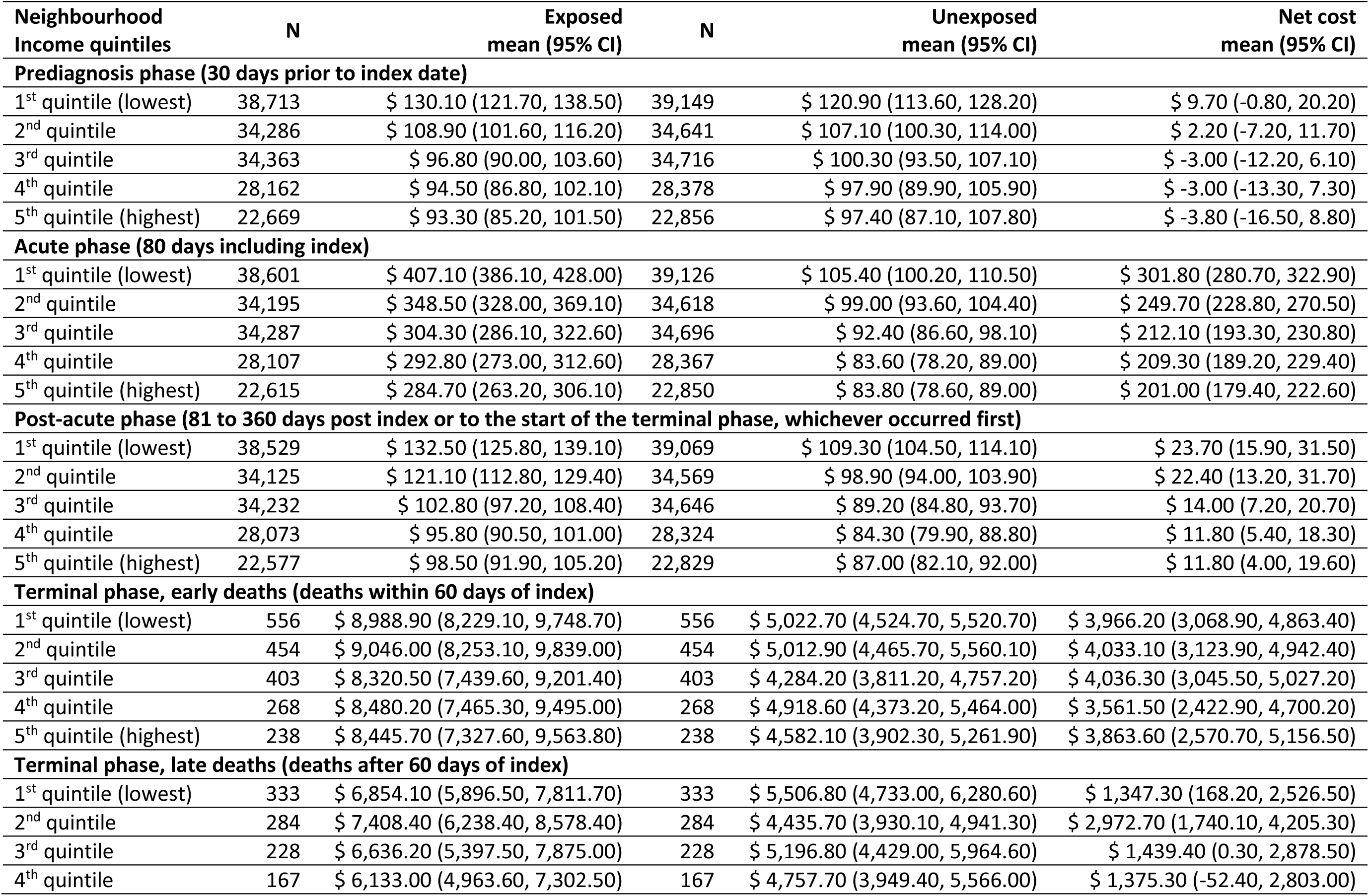
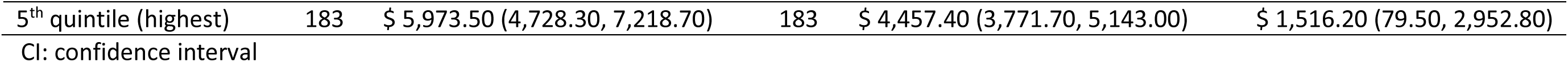
Total and COVID-19-attributable healthcare costs (2023 CAD) standardized to 10 days stratified by neighbourhood income quintile.

**Table S6.** Total and COVID-19-attributable healthcare costs (2023 CAD) standardized to 10 days stratified by resource utilization band.

| RUB | N | Exposed<br>mean (95% CI) | N | Unexposed<br>mean (95% CI) | Net cost<br>mean (95% CI) |
| --- | --- | --- | --- | --- | --- |
| **Prediagnosis phase (30 days prior to index date)** |  |  |  |  |  |
| Non-users | 12,270 | \$ 3.20 (2.90, 3.50) | 12,292 | \$ 3.40 (3.20, 3.60) | \$ -0.20 (-0.60, 0.20) |
| Healthy Users | 8,130 | \$ 7.40 (7.00, 7.80) | 8,138 | \$ 8.50 (7.90, 9.10) | \$ -1.10 (-1.80, -0.40) |
| Low resource utilization | 28,086 | \$ 15.70 (14.90, 16.60) | 28,131 | \$ 17.60 (16.60, 18.70) | \$ -1.90 (-3.30, -0.50) |
| Moderate resource utilization | 77,704 | \$ 53.60 (51.70, 55.50) | 78,141 | \$ 57.80 (55.80, 59.80) | \$ -4.20 (-6.90, -1.50) |
| High resource utilization | 23,664 | \$ 176.40 (167.40, 185.40) | 24,023 | \$ 189.10 (179.50, 198.60) | \$ -12.60 (-25.50, 0.20) |
| Very high resource utilization | 8,339 | \$ 958.60 (903.80, 1,013.40) | 9,015 | \$ 805.90 (755.80, 856.00) | \$ 153.30 (80.30, 226.30) |
| **Acute phase (79 days including index)** |  |  |  |  |  |
| Non-users | 12,268 | \$ 110.80 (88.70, 132.90) | 12,291 | \$ 6.90 (6.10, 7.60) | \$ 103.90 (81.80, 126.10) |
| Healthy Users | 8,127 | \$ 76.80 (54.40, 99.20) | 8,137 | \$ 12.10 (11.00, 13.20) | \$ 64.70 (42.30, 87.10) |
| Low resource utilization | 28,068 | \$ 112.60 (99.50, 125.70) | 28,131 | \$ 20.70 (19.50, 22.00) | \$ 91.90 (78.70, 105.10) |
| Moderate resource utilization | 77,571 | \$ 276.00 (263.70, 288.30) | 78,131 | \$ 59.30 (57.30, 61.20) | \$ 216.70 (204.30, 229.20) |
| High resource utilization | 23,581 | \$ 486.30 (461.70, 511.00) | 24,012 | \$ 166.40 (158.60, 174.10) | \$ 320.00 (294.40, 345.60) |
| Very high resource utilization | 8,190 | \$ 1,796.10 (1,708.80, 1,883.40) | 8,955 | \$ 630.70 (598.70, 662.80) | \$ 1,165.70 (1,073.90, 1,257.50) |
| **Post-acute phase (81 to 360 days post index or to the start of the terminal phase, whichever occurred first)** |  |  |  |  |  |
| Non-users | 12,264 | \$ 23.60 (19.90, 27.30) | 12,291 | \$ 13.40 (11.20, 15.60) | \$ 10.20 (5.90, 14.50) |
| Healthy Users | 8,126 | \$ 21.70 (15.40, 28.10) | 8,134 | \$ 17.70 (14.40, 21.10) | \$ 4.00 (-3.20, 11.20) |
| Low resource utilization | 28,062 | \$ 30.40 (28.10, 32.60) | 28,130 | \$ 26.40 (24.80, 27.90) | \$ 4.00 (1.30, 6.70) |
| Moderate resource utilization | 77,515 | \$ 79.60 (76.40, 82.80) | 78,092 | \$ 66.10 (64.10, 68.10) | \$ 13.50 (9.80, 17.20) |
| High resource utilization | 23,530 | \$ 178.40 (170.10, 186.60) | 23,966 | \$ 159.60 (153.10, 166.10) | \$ 18.90 (8.50, 29.20) |
| Very high resource utilization | 8,039 | \$ 743.80 (703.50, 784.10) | 8,824 | \$ 580.10 (553.20, 607.10) | \$ 164.40 (117.10, 211.60) |
| **Terminal phase, early deaths (deaths within 60 days of index)** |  |  |  |  |  |
| Non-users | 18 | \$ 9,762.40 (4,637.90, 14,887.00) | 18 | \$ 6,109.10 (1,262.40, 10,955.80) | \$ 3,653.30 (-2,943.70, 10,250.30) |
| Healthy Users | 7 | \$ 13,280.20 (3,054.30, 23,506.20) | 7 | \$ 2,008.90 (-452.10, 4,469.90) | \$ 11,271.40 (3,288.30, 19,254.40) |
| Low resource utilization | 56 | \$ 11,253.20 (8,758.80, 13,747.60) | 56 | \$ 3,759.90 (2,482.80, 5,037.00) | \$ 7,493.30 (4,700.90, 10,285.70) |
| Moderate resource utilization | 553 | \$ 9,422.80 (8,582.60, 10,262.90) | 553 | \$ 4,607.40 (4,151.30, 5,063.50) | \$ 4,815.40 (3,898.50, 5,732.30) |
| High resource utilization | 427 | \$ 8,589.40 (7,731.40, 9,447.40) | 427 | \$ 4,444.80 (3,942.60, 4,947.10) | \$ 4,144.50 (3,176.50, 5,112.60) |
| Very high resource utilization | 858 | \$ 8,115.70 (7,600.70, 8,630.70) | 858 | \$ 5,155.40 (4,786.50, 5,524.20) | \$ 2,960.40 (2,334.80, 3,585.90) |
| **Terminal phase, late deaths (deaths after 60 days of index)** |  |  |  |  |  |
| Non-users | 17 | \$ 9,953.60 (3,943.20, 15,963.90) | 17 | \$ 2,194.80 (529.40, 3,860.10) | \$ 7,758.80 (2,175.00, 13,342.60) |
| Healthy Users | <=5 | \$ 6,862.00 (-11,617.90, 25,341.90) | <=5 | \$ 4,340.00 (-6,166.90, 14,846.90) | \$ 2,522.00 (-8,245.50, 13,289.40) |
| Low resource utilization | 23 | \$ 5,543.40 (2,198.10, 8,888.60) | 23 | \$ 3,596.30 (1,429.20, 5,763.40) | \$ 1,947.00 (-1,987.40, 5,881.50) |
| Moderate resource utilization | 279 | \$ 7,754.90 (6,278.30, 9,231.40) | 279 | \$ 4,928.70 (4,202.00, 5,655.30) | \$ 2,826.20 (1,201.40, 4,451.00) |
| High resource utilization | 231 | \$ 6,276.20 (5,133.40, 7,418.90) | 231 | \$ 4,040.00 (3,434.50, 4,645.50) | \$ 2,236.10 (947.30, 3,524.90) |
| Very high resource utilization | 642 | \$ 6,364.70 (5,812.20, 6,917.30) | 642 | \$ 5,369.50 (4,911.80, 5,827.30) | \$ 995.20 (307.70, 1,682.60) |

CI: confidence interval; RUB: resource utilization band

**Table S7.** Total and COVID-19-attributable healthcare costs (2023 CAD) standardized to 10 days after varying pre-diagnosis, acute and post-acute phase lengths.

| Costs categories | Exposed<br>mean (95% CI)<br>N=159,817 | Unexposed<br>mean (95% CI)<br>N=159,817 | Net cost<br>mean (95% CI)<br>N=159,817 |
| --- | --- | --- | --- |
| **Prediagnosis phase (120 days prior to index date)** |  |  |  |
| Total costs | \$ 102.70 (100.00, 105.40) | \$ 97.20 (94.80, 99.60) | \$ 5.50 (2.20, 8.90) |
| Costs by resource category |  |  |  |
| Inpatient care | \$ 25.00 (23.40, 26.70) | \$ 23.90 (22.30, 25.40) | \$ 1.20 (-1.00, 3.30) |
| Outpatient services | \$ 16.90 (16.20, 17.70) | \$ 14.60 (14.10, 15.20) | \$ 2.30 (1.40, 3.20) |
| ED visits | \$ 5.00 (4.90, 5.10) | \$ 3.70 (3.60, 3.80) | \$ 1.30 (1.20, 1.40) |
| Medications | \$ 12.20 (11.80, 12.70) | \$ 11.40 (10.90, 11.90) | \$ 0.90 (0.20, 1.50) |
| Physician services | \$ 4.90 (4.80, 5.00) | \$ 5.20 (5.10, 5.20) | \$ -0.30 (-0.30, -0.20) |
| Rehabilitation | \$ 2.10 (1.70, 2.50) | \$ 2.00 (1.70, 2.30) | \$ 0.10 (-0.40, 0.60) |
| Complex care | \$ 3.50 (2.80, 4.20) | \$ 2.80 (2.20, 3.40) | \$ 0.70 (-0.20, 1.60) |
| Homecare | \$ 8.80 (8.30, 9.30) | \$ 6.90 (6.60, 7.30) | \$ 1.90 (1.30, 2.50) |
| Long term care | \$ 0.00 (,,) | \$ 0.00 (,,) | \$ 0.00 (,,) |
| Other | \$ 1.90 (1.80, 1.90) | \$ 2.80 (2.80, 2.80) | \$ -0.90 (-1.00, -0.90) |
| **Acute phase (29 days including index)** |  |  |  |
| Total costs | \$ 556.10 (542.20, 570.00) | \$ 99.70 (96.70, 102.70) | \$ 456.50 (442.50, 470.50) |
| Costs by resource category |  |  |  |
| Inpatient care | \$ 378.90 (366.80, 391.00) | \$ 21.80 (19.90, 23.70) | \$ 357.10 (344.90, 369.30) |
| Outpatient services | \$ 19.40 (18.60, 20.10) | \$ 14.90 (14.20, 15.60) | \$ 4.50 (3.50, 5.50) |
| ED visits | \$ 39.90 (39.40, 40.50) | \$ 3.70 (3.60, 3.80) | \$ 36.20 (35.60, 36.80) |
| Medications | \$ 10.90 (10.40, 11.40) | \$ 11.90 (11.30, 12.50) | \$ -1.00 (-1.80, -0.20) |
| Physician services | \$ 13.90 (13.70, 14.00) | \$ 7.50 (7.40, 7.50) | \$ 6.40 (6.20, 6.60) |
| Rehabilitation | \$ 6.20 (5.10, 7.20) | \$ 1.90 (1.40, 2.50) | \$ 4.20 (3.10, 5.40) |
| Complex care | \$ 4.60 (3.70, 5.40) | \$ 3.40 (2.70, 4.20) | \$ 1.20 (0.10, 2.30) |
| Homecare | \$ 11.10 (10.60, 11.70) | \$ 8.10 (7.60, 8.50) | \$ 3.20 (2.50, 3.80) |
| Long term care | \$ 0.10 (0.00, 0.10) | \$ 0.20 (0.10, 0.30) | \$ -0.20 (-0.30, 0.00) |
| Other | \$ 1.10 (1.10, 1.10) | \$ 2.70 (2.70, 2.80) | \$ -1.60 (-1.70, -1.60) |
| **Post-acute phase (81 to 360 days post index or to the start of the terminal phase, whichever occurred first)** |  |  |  |
| Total costs | \$ 130.70 (126.80, 134.60) | \$ 95.60 (93.40, 97.70) | \$ 35.40 (31.10, 39.70) |
| Costs by resource category |  |  |  |
| Inpatient care | \$ 44.10 (41.10, 47.10) | \$ 22.30 (21.00, 23.60) | \$ 21.90 (18.60, 25.10) |
| Outpatient services | \$ 18.20 (17.50, 18.90) | \$ 14.90 (14.40, 15.50) | \$ 3.30 (2.40, 4.20) |
| ED visits | \$ 5.00 (4.90, 5.10) | \$ 3.60 (3.50, 3.60) | \$ 1.40 (1.30, 1.60) |
| Medications | \$ 12.10 (11.70, 12.50) | \$ 11.80 (11.30, 12.20) | \$ 0.30 (-0.30, 0.90) |
| Physician services | \$ 4.60 (4.50, 4.60) | \$ 4.60 (4.60, 4.70) | \$ -0.10 (-0.10, 0.00) |
| Rehabilitation | \$ 2.90 (2.60, 3.30) | \$ 1.40 (1.20, 1.70) | \$ 1.50 (1.10, 1.90) |
| Complex care | \$ 5.30 (4.60, 6.00) | \$ 3.00 (2.50, 3.60) | \$ 2.30 (1.40, 3.20) |
| Homecare | \$ 8.50 (8.10, 9.00) | \$ 7.10 (6.70, 7.40) | \$ 1.50 (1.00, 2.10) |
| Long term care | \$ 4.00 (3.50, 4.40) | \$ 2.70 (2.40, 3.10) | \$ 1.20 ( 0.70, 1.80) |
| Other | \$ 2.00 (2.00, 2.00) | \$ 2.50 (2.50, 2.60) | \$ -0.50 (-0.60, -0.50) |

ED: emergency department; CI: confidence interval

**Table S8.** Total and COVID-19-attributable healthcare costs (2023 CAD), stratified by hospitalization and mortality.

| **Total Cost during overall** | **Matched Exposed, N** | **Matched Exposed, 10d<sup>1</sup> mean (95%CI) costs** | **Attributable Costs 10d<sup>1</sup> mean (95%CI)** | **Matched Exposed, Total<sup>2</sup> mean (95%CI) costs** | **Attributable Costs Total<sup>2</sup> mean (95%CI)** |
| --- | --- | --- | --- | --- | --- |
| No hospital stay within 14 days of index | 151,716 | \$ 119<br>(114; 124) | \$ 26<br>(20; 31) | \$ 3,282<br>(3,211; 3,353) | \$ 295<br>(207; 383) |
| Hospital stay within 14 days of index | 8,101 | \$ 6,905<br>(6,627; 7,183) | \$ 6,499<br>(6,221; 6,777) | \$ 61,384<br>(59,589; 63,179) | \$ 50,490<br>(48,608; 52,372) |
| **Of those with a hospitalization within 14 days** |  |  |  |  |  |
| No ICU stay | 5,950 | \$ 3,610<br>(3,436; 3,784) | \$ 3,174<br>(3,000; 3,348) | \$ 41,522<br>(40,280; 42,764) | \$ 30,147<br>(28,760; 31,534) |
| ICU stay within 14 days of admission | 2,126 | \$ 15,960<br>(15,140; 16,780) | \$ 15,636<br>(14,818; 16,454) | \$ 115,271<br>(110,181; 120,361) | \$ 105,677<br>(100,467; 110,888) |
| ICU stay after 14 days of admission | 25 | \$ 21,163<br>(13,537; 28,788) | \$ 20,935<br>(13,902; 27,969) | \$ 205,842<br>(108,022; 303,661) | \$ 198,971<br>(107,437; 290,506) |
| **Of those with a hospitalization within 14 days** |  |  |  |  |  |
| No ICU stay, alive at end of follow up | 4,757 | \$ 1,083<br>(1,047; 1,119) | \$ 746<br>(696; 796) | \$ 38,987<br>(37,707; 40,267) | \$ 30,030<br>(28,672; 31,389) |
| No ICU stay, died before end of follow up | 1,193 | \$ 13,687<br>(13,118; 14,256) | \$ 12,854<br>(12,275; 13,433) | \$ 51,631<br>(48,180; 55,083) | \$ 30,610<br>(26,310; 34,911) |
| ICU stay within 14 days of admission, alive at end of follow up | 1,300 | \$ 3,410<br>(3,221; 3,598) | \$ 3,164<br>(2,971; 3,357) | \$ 122,744<br>(115,957; 129,532) | \$ 114,945<br>(108,101; 121,790) |
| ICU stay within 14 days of admission, died before end of follow up | 826 | \$ 35,712<br>(34,529; 36,894) | \$ 35,266<br>(34,078; 36,453) | \$ 103,509<br>(95,976; 111,042) | \$ 91,091<br>(83,205; 98,977) |
| ICU stay after 14 days of admission, alive at end of follow up | 10 | \$ 6,231<br>(2,270; 10,193) | \$ 6,160<br>(2,883; 9,437) | \$ 224,325<br>(81,710; 366,940) | \$ 221,754<br>(103,788; 339,720) |
| ICU stay after 14 days of admission, died before end of follow up | 15 | \$ 31,117<br>(21,527; 40,707) | \$ 30,786<br>(22,388; 39,183) | \$ 193,519<br>(46,675; 340,364) | \$ 183,783<br>(53,625; 313,941) |

CAD: Canadian dollar, CI: confidence interval; d: day; ICU: intensive care unit
<sup>1</sup> Mean 10 day costs, rounded to the nearest dollar, for the period from index to end of follow-up or death whichever occurred first.
<sup>2</sup> Mean total costs, rounded to the nearest dollar, from index to end of follow-up or death whichever occurred first.

## References

[1] Canadian Institute for Health Information. Impact of COVID-19 on Canada’s health care systems (2023). Available at https://www.cihi.ca/en/covid-19-resources/impact-of-covid-19-on-canadas-health-care-systems Accessed on June 17, 2024.

[2] Quinn KL, Katz GM, Bobos P, Sander B, McNaughton CD, Cheung AM, et al. Understanding the post COVID-19 condition (long COVID) in adults and the expected burden for Ontario. Science Briefs of the Ontario COVID-19 Science Advisory Table. 2022;3(65).

[3] Katz GM, Bach K, Bobos P, Cheung A, Decary S, Goulding S, et al. Understanding How Post-COVID-19 Condition Affects Adults and Health Care Systems. JAMA Health Forum. 2023;4(7):e231933.

[4] World Health Organisation. Post COVID-19 condition, Long COVID (2022). Available at https://www.who.int/europe/news-room/fact-sheets/item/post-covid-19-condition Accessed on June 17, 2024

[5] Blanchflower DG, Bryson A. Long COVID in the United States. PLoS One. 2023;18(11):e0292672.

[6] Government of Canada. PostCovid-19 condition (long COVID). Available at https://www.canada.ca/en/public-health/services/diseases/2019-novel-coronavirus-infection/symptoms/post-covid-19-condition.html Accessed on July 24, 2024.

[7] McNaughton CD, Austin PC, Sivaswamy A, Fang J, Abdel-Qadir H, Daneman N, et al. Post-acute health care burden after SARS-CoV-2 infection: a retrospective cohort study. CMAJ. 2022;194(40):E1368–E76.

[8] Tartof SY, Malden DE, Liu IA, Sy LS, Lewin BJ, Williams JTB, et al. Health Care Utilization in the 6 Months Following SARS-CoV-2 Infection. JAMA Netw Open. 2022;5(8):e2225657.

[9] Whittaker HR, Gulea C, Koteci A, Kallis C, Morgan AD, Iwundu C, et al. GP consultation rates for sequelae after acute covid-19 in patients managed in the community or hospital in the UK: population based study. BMJ. 2021;375:e065834.

[10] Patterson B, Ruppenkamp J, Richards F, Debnath R, ElKhoury AC, DeMartino JK, et al. Cost of long COVID following severe diseasea US healthcare database analysis. Value Health. 2022;25(7):S375.

[11] The COVID-19 Immunity Task Force. Seroprevalence in Canada. Available at https://www.covid19immunitytaskforce.ca/seroprevalence-in-canada/ Accessed on June 17, 2024.

[12] Shrestha SS, Kompaniyets L, Grosse SD, Harris AM, Baggs J, Sircar K, et al. Estimation of Coronavirus Disease 2019 Hospitalization Costs From a Large Electronic Administrative Discharge Database, March 2020-July 2021. Open Forum Infect Dis. 2021;8(12):ofab561.

[13] Yang J, Andersen KM, Rai KK, Tritton T, Mugwagwa T, Reimbaeva M, et al. Healthcare resource utilisation and costs of hospitalisation and primary care among adults with COVID-19 in England: a population-based cohort study. BMJ Open. 2023;13(12):e075495.

[14] Sagy WY, Feldhamer I, Brammli-Greenberg S, Lavie G. Estimating the economic burden of long-Covid: the additive cost of healthcare utilisation among COVID-19 recoverees in Israel. BMJ Glob Health. 2023;8(7).

[15] McNaughton CD, Austin PC, Li Z, Sivaswamy A, Fang J, Abdel-Qadir H, et al. Post-acute health care costs following SARS-CoV-2 infection: A retrospective cohort study of among 531,182 matched adults. medRxiv the preprint server for health sciences.

[16] Benchimol EI, Smeeth L, Guttmann A, Harron K, Moher D, Petersen I, et al. The REporting of studies Conducted using Observational Routinely-collected health Data (RECORD) statement. PLoS Med. 2015;12(10):e1001885.

[17] Government of Canada. COVID-19 vaccination: Doses administered. Available at https://health-infobase.canada.ca/covid-19/vaccine-administration/ Accessed on June 17, 2024.

[18] Dave N, Sjoholm D, Hedberg P, Ternhag A, Granath F, Verberk JDM, et al. Nosocomial SARS-CoV-2 Infections and Mortality During Unique COVID-19 Epidemic Waves. JAMA Netw Open. 2023;6(11):e2341936.

[19] Mitchell R, Choi KB, Pelude L, Rudnick W, Thampi N, Taylor G, et al. Patients in hospital with laboratory-confirmed COVID-19 in a network of Canadian acute care hospitals, Mar. 1 to Aug. 31, 2020: a descriptive analysis. CMAJ Open. 2021;9(1):E149–E56.

[20] Public Health Ontario. Best Practices for Managing COVID-19 Outbreaks in Acute Care Settings 2nd edition: January 2023. Available at https://www.publichealthontario.ca/-/media/Documents/nCoV/ipac/2021/03/covid-19-pidac-outbreaks-acute-care.pdf.

[21] Zeitouny S, Cheung DC, Bremner KE, Pataky RE, Pequeno P, Matelski J, et al. The impact of the early COVID-19 pandemic on healthcare system resource use and costs in two provinces in Canada: An interrupted time series analysis. PLoS One. 2023;18(9):e0290646.

[22] Canadian Institute for Health Information. Canadian COVID-19 Intervention Timeline. Available at https://www.cihi.ca/en/canadian-covid-19-intervention-timeline Accessed on June 25, 2024.

[23] Glazier RH, Green ME, Wu FC, Frymire E, Kopp A, Kiran T. Shifts in office and virtual primary care during the early COVID-19 pandemic in Ontario, Canada. CMAJ. 2021;193(6):E200–E10.

[24] Public Health Ontario. The Story of COVID-19 Testing in Ontario. Available at https://www.publichealthontario.ca/en/About/News/2020/Story-Covid-19-Testing-Ontario Accessed on June 25, 2024.

[25] Action to beat coronavirus/Action pour battre le coronavirus (Ab-C) Study Investigators Jha P. COVID Seroprevalence, Symptoms and Mortality During the First Wave of SARS-CoV-2 in Canada. medRxiv. 2021 DOI: 10.1101/2021.03.04.21252540.

[26] Austin PC. An Introduction to Propensity Score Methods for Reducing the Effects of Confounding in Observational Studies. Multivariate Behav Res. 2011;46(3):399–424.

[27] Austin PC. Optimal caliper widths for propensity-score matching when estimating differences in means and differences in proportions in observational studies. Pharm Stat. 2011;10(2):150–61.

[28] The Johns Hopkins University. The Johns Hopkins ACG System. Excerpt from Version11.0 Technical Reference Guide. November 2014. Available at https://www2.gov.bc.ca/assets/gov/health/conducting-health-research/data-access/johns-hopkins-acg-system-technical-reference-guide.pdf Accessed on July 24, 2024.

[29] Johns Hopkins Medicine. The ACG System is an essential tool in helping providers manage multiple chronic conditions. Available at https://www.hopkinsacg.org/the-acg-system-is-an-essential-tool-in-helping-providers-manage-multiple-chronic-conditions/ Accessed on July 24, 2024.

[30] Matheson FI, Moloney G, van Ingen T, Joint publication with Public Health Ontario. Ontario marginalization index: user guide (2021). Toronto, ON: St. Michael’s Hospital (Unity Health Toronto); 2023.

[31] Johns Hopkins Medicine. Johns Hopkins ACG® System version 13.0. Available at https://www.hopkinsacg.org/ Accessed on December 24, 2022.

[32] Tanuseputro P, Wodchis WP, Fowler R, Walker P, Bai YQ, Bronskill SE, et al. The health care cost of dying: a population-based retrospective cohort study of the last year of life in Ontario, Canada. PLoS One. 2015;10(3):e0121759.

[33] Wodchis WP, Bushmeneva K, Nikitovic M, Mckillop I. Guidelines on Person-Level Costing Using Administrative Databases in Ontario. Available: http://www.hsprn.ca/uploads/files/Guidelines_on_PersonLevel_Costing_May_2013.pdf. 2013.

[34] Yabroff KR, Lamont EB, Mariotto A, Warren JL, Topor M, Meekins A, et al. Cost of care for elderly cancer patients in the United States. J Natl Cancer Inst. 2008;100(9):630–41.

[35] Kim HJ, Fay MP, Feuer EJ, Midthune DN. Permutation tests for joinpoint regression with applications to cancer rates. Stat Med. 2000;19(3):335–51.

[36] Parotto M, Gyongyosi M, Howe K, Myatra SN, Ranzani O, Shankar-Hari M, et al. Post-acute sequelae of COVID-19: understanding and addressing the burden of multisystem manifestations. Lancet Respir Med. 2023;11(8):739–54.

[37] World Health organisation. A clinical case definition of post COVID-19 condition by a Delphi consensus, 6 October 2021. Available at https://www.who.int/publications/i/item/WHO-2019-nCoV-Post_COVID-19_condition-Clinical_case_definition-2021.1 Accessed on June, 14, 2024.

[38] Soriano JB, Murthy S, Marshall JC, Relan P, Diaz JV, Condition WHOCCDWGoP-C-. A clinical case definition of post-COVID-19 condition by a Delphi consensus. Lancet Infect Dis. 2022;22(4):e102–e7.

[39] Centers for Disease Control and Prevention. Clinical Overview of Long Covid. Available at https://www.cdc.gov/coronavirus/2019-ncov/hcp/clinical-care/post-covid-conditions.html Accessed on June, 24, 2024.

[40] Tsui TCO, Zeitouny S, Bremner KE, Cheung DC, Mulder C, Croxford R, et al. Initial health care costs for COVID-19 in British Columbia and Ontario, Canada: an interprovincial population-based cohort study. CMAJ Open. 2022;10(3):E818–E30.

[41] Wang L, Calzavara A, Baral S, Smylie J, Chan A.K., Sander B, Austin PC, Kwong JC, Mishra S. Differential Patterns by Area-Level Social Determinants of Health in Coronavirus Disease 2019 (COVID-19)–Related Mortality and Non–COVID-19 Mortality: A Population-Based Study of 11.8 Million People in Ontario, Canada, Clinical Infectious Diseases. 2023;76(6):1110–1120, 10.1093/cid/ciac850

